# A 3D ovarian cancer metastasis model using a decellularized peritoneal matrix to study therapy response

**DOI:** 10.1101/2024.12.27.24319681

**Authors:** Christiane Helgestad Gjerde, Katrin Kleinmanns, Anika Langer, Gorka Ruiz de Garibay Ponce, Ezekiel Rozmus, Gina Nyhus Stangeland, Calum Leitch, Rammah Elnour, Harsh Nitin Dongre, Constantin Berger, Okan Gultekin, Christopher Forcados, Maria Stensland, Tuula A. Nyman, Kaisa Lehti, Ben Davidson, Sébastien Wälchli, Pascal Gelebart, Daniela Elena Costea, Spiros Kotopoulis, Line Bjørge, Emmet McCormack

## Abstract

High-grade serous ovarian carcinoma (HGSOC) presents a significant therapeutic challenge. Late-stage disease is frequently associated with peritoneal carcinomatosis. The peritoneal metastases exhibit a unique tumor microenvironment (TME) distinct from the primary tumors and other metastatic sites. Understanding the critical influence of the extracellular matrix (ECM) in shaping the tumor phenotype is essential for the development of effective new therapies. This study introduces a novel three-dimensional (3D) model of HGSOC peritoneal metastases using a porcine decellularized peritoneal-derived ECM scaffold, referred to as peritoneal matrix (PerMa). We show that the decellularization maintains the structural integrity and composition of ECM molecules. Comparative analysis reveals structural, compositional, and mechanical similarities between porcine and human peritoneal matrices, underscoring the porcine model’s translational relevance for modeling human peritoneum physiology. The PerMa supports the 3D growth of HGSOC cell lines. The model enables the assessment of sensitivity to traditional chemotherapy and novel cell-based immunotherapy through confocal imaging and quantification of cell volume. Our model offers a valuable platform for investigating peritoneal carcinomatosis in HGSOC, with the potential to contribute significantly to the development of novel therapeutic approaches.

## 1 Introduction

Epithelial ovarian cancer (EOC) is the most lethal gynecological malignancy with an estimated 310,000 new cases and 200,000 new deaths every year (1). The prognosis for patients heavily depends on the stage at diagnosis, with most being diagnosed at a late stage, resulting in a 5-year survival rate of less than 30% for this group (2, 3). High-grade serous ovarian carcinoma (HGSOC) is the most aggressive subtype of EOC, accounting for the majority of EOC-related deaths (4, 5). HGSOC primarily spreads across the peritoneal cavity, referred to as transcoelomic metastasis, forming metastatic deposits on the mesothelial lining of the cavity (6, 7). The peritoneum, a specialized serous membrane, lines the abdominal walls (peritoneum parietale) and internal organs (peritoneum viscerale) (8). The malignant cells follow the dynamics of the peritoneal fluid and preferentially distribute to areas with constant exposure to peritoneal fluid (the omentum) and areas where fluid accumulates (the Pouch of Douglas and the right subphrenic region). This pattern of spread leads to peritoneal carcinomatosis, marked by widespread and diffuse metastases that significantly complicate surgical resection (9).

Metastases on the visceral and parietal peritoneum are typically numerous, small, and superficial, with distinct phenotypic features compared to omental metastases and other metastatic sites (10–13). Examining HGSOC transcriptome and histology, Pietilä et al. describe the fibrotic ECM heterogeneity at ovarian tumors and other metastatic sites, including the peritoneum and the omentum in treatment naïve and treated HGSOCs. These results also demonstrate that the desmoplastic ECM signaling enhances HGSOC chemoresistance by ECM composition- and stiffness-induced β1integrin-FAK-pMLC-YAP signaling (13). A recent study by Vázquez-García et al. underscored how differences in the local microenvironment at various metastatic sites influence the formation of the tumor microenvironment (TME) (11). They observed disparities in tumor-immune cell interactions between HGSOC ovarian tumors and metastases from different anatomical sites, such as bowel and adipose tissue. How the different cellular and non-cellular components of the local TME impact these interactions remains to be elucidated. The extracellular matrix (ECM) plays an important role in disease aggressiveness and therapy responses (14, 15). The ECM is a macromolecular network of proteins and polysaccharides that provides physical scaffolding for cells and regulates cell differentiation, proliferation, migration, and drug resistance (16). In the metastatic process, HGSOC cells adhere to the peritoneal lining and invade the underlying stroma (17). They contribute to ECM remodeling through interaction with peritoneal cells, which promotes metastasis (18). ECM remodeling during tumor progression and in response to chemotherapy also affects immune cell infiltration (19, 20), which in turn can influence the response to immunotherapeutics (21).

Given that it is the metastatic disease that ultimately leads to high patient mortality in HGSOC, improving our understanding of adhesion, invasion, and ECM remodeling in HGSOC metastases is crucial. To achieve this, we need advanced models that accurately replicate the complexity of the different metastatic sites, for better disease modeling and drug testing. Three-dimensional (3D) *in vitro* models recapitulate different aspects of tumor biology. Patient-derived organoids (PDOs) have been shown to preserve the genomic features of the original patient tumor (22–26). However, 3D cultures based on patient-derived cancer stem cells fail to model the cell-cell and cell-matrix interactions within the TME (27). On the other hand, multicellular 3D organotypic models more accurately mimic the heterogenous *in vivo* tissue architecture (27). Despite this improvement, most models still rely on hydrogels, such as collagen I gels or Matrigel®, which inadequately replicate the complex ECM of tissues. The patient-derived omental matrix gel (OmGel) mimics the HGSOC omental metastasis protein composition and is shown to be a promising tool for studying the biology of omental metastases (28). However, the complex architecture of the ECM is not well preserved in hydrogels. Biological matrices generated by tissue decellularization offer a promising alternative with a preserved ECM structure and biomolecules critical for cell behavior and response to therapy (29). Small intestinal submucosa (SIS) derived from pigs has been used to generate 3D models of colorectal cancer (30) and more recently EOC (31). Considering the heterogeneity in the interconnected ECM and immune TME across metastatic sites (11, 13), experimental models that accurately mimic the specific ECM environments are of critical need to study and target the mechanisms of HGSOC metastasis and therapy response and resistance. Thus, a peritoneal decellularized ECM (dECM) scaffold would be preferred to model peritoneal carcinomatosis.

In this study, we developed a novel dECM scaffold referred to as a peritoneal matrix (PerMa) from porcine and human parietal peritoneum to better mimic the peritoneal ECM. The decellularization effectively removed cells and genetic material while maintaining the structural integrity of major ECM components. Due to the observed resemblance to the human peritoneum in this study, the porcine PerMa (pPerMa) was used, offering advantages as a scaffold for 3D culture in terms of availability and reproducibility. The peritoneal scaffold supported the 3D growth of HGSOC cell lines. We further demonstrated the utility of the model for assessing sensitivity to traditional chemotherapy and novel, cell-based immunotherapy. The 3D PerMa model is a promising platform for studying the complex interplay between HGSOC cells and the peritoneal microenvironment and how the interaction affects therapy sensitivity.

## 2 Materials and methods

### 2.1 Materials

#### 2.1.1 Cell lines and reagents

Human EOC cell lines Caov3 (cat# HTB-75, RRID: CVCL_0201), OV90 (cat# CRL-11732, RRID: CVCL_3768), and OVCAR3 (cat # HTB-161, RRID: CVCL_0465) were obtained from the American Type Culture Collection (ATCC Manassas, VA, USA), and human EOC cell line COV318 (cat# 07071903, RRID: CVCL_2419) from Sigma Aldrich (Sigma Aldrich, St. Louis, MO, USA). The human fibroblast cell line BJ was kindly provided by Professor Donald Gullberg (Department of Biomedicine, University of Bergen, Norway).

The cell lines were cultured in RPMI 1640 (OV-90) or DMEM high glucose (Caov-3, COV318, BJ fibroblasts) media supplemented with 10% heat-inactivated fetal bovine serum (FBS), 1% L-glutamine and 1% penicillin-streptomycin (P/S) (RPMI cat# R5886, DMEM cat# D5671, penicillin-streptomycin cat# P0781, Sigma Aldrich; FBS cat# 10270106, L-Glutamine cat# 25030081, Gibco, Waltham, MA, USA). For co-culture experiments with OV90 and BJ fibroblasts, the cells were cultured in a 1:1 ratio of RPMI/DMEM.

Human EOC cell lines Caov-3, OV-90, and COV318 were transduced to express green fluorescent protein (GFP), performed according to the manufacturer’s protocol using the Redi-Fect Red-FLuc-GFP lentiviral particles (cat# CLS960003, Perkin Elmer, Waltham, MA, USA). The BJ fibroblast cell line was transduced to express miRFP670, performed according to the manufacturers protocol using the pLenti6.2_miRFP670 lentiviral particles (pLenti6.2_miRFP670 was a gift from Vanessa LaPointe, Addgene plasmid# 113726; http://n2t.net/addgene:113726 ; RRID: Addgene_113726).

#### 2.1.2 Human and animal tissues

Animal experiments were performed in accordance with the procedures from the Norwegian Commission for Laboratory Animals, the European Convention for the Protection of Vertebrates Used for Scientific Purposes (EU Directive 2010/63/EU for animal experiments), and the ethical standards given in the Declaration of Helsinki of 2008.

Porcine small intestine and peritoneum were surplus material from animals used for surgical training or in research at the Laboratory Animal Facility, Department of Clinical Medicine, University of Bergen, Norway. The samples were collected shortly post-mortem, within 1 hour. The surgical training and the research experiments in animals were approved by the Norwegian Food Safety Authority (FOTS IDs: 29559, 13911, 13904, 10294, 20330, and 21065). According to The Norwegian State Commission for Laboratory Animals, there was no need for additional approvals from the Norwegian Food Safety Authority for the use of remnant material from the dead test animals.

Human healthy peritoneum was obtained from autopsies of women at the Department of Pathology, Haukeland University Hospital, Bergen, Norway. Informed consent was obtained from the patient’s closest relative before the collection of material. Women with intra-abdominal inflammation, cancer, or peritoneal carcinomatosis were excluded from the study. The material was collected within 48 hours after death.

Peritoneum from ovarian cancer patients was collected from chemotherapy naïve patients with primary advanced high-grade serous ovarian carcinoma (HGSOC), admitted to the Department of Obstetrics and Gynecology, Haukeland University Hospital, Bergen, Norway. The peritoneum was collected from a region with no macroscopically visible metastasis. The peritoneal tissue specimens were included in the Bergen Gynecologic Cancer Biobank. Informed consent was obtained from the women before collection of the samples.

The regional committees for Medical and Health Research Ethics (REK West) approved the biobank and the studies (Reference IDs: 2014/1907, 30477).

### 2.2 Generation of SIS from porcine intestines

Production of small intestinal submucosa matrix (SIS) by decellularization of porcine jejunum was performed as described previously (32). Briefly, segments of jejunum were cleaned with tap water and cut into 20 cm segments. Intestinal segments were inverted, and mucosa was scraped off using forceps, followed by incubation overnight in Ca^2+^- and Mg^2+^-free phosphate buffered saline (PBS^—^) with 1% P/S. Prior to decellularization, specimens for DNA quantification, proteomic and histopathological analysis were collected. Samples for histopathological analysis were fixed in 4% paraformaldehyde (PFA) for 1 hour at room temperature and paraffin-embedded. Samples for DNA quantification and proteomics were frozen at −80 °C. For tissue decellularization, intestinal segments were first filled with 3.4% sodium deoxycholate (DOC; cat # D6750, Sigma Aldrich, St. Louis, USA) dissolved in Milli-Q (MQ) water and incubated at 4 °C for 1.5 hours with a magnetic stirrer. After washing 5 times during 1 hour with PBS^—^ containing 1% P/S, SIS pieces were stored in PBS^—^ containing 1% P/S overnight. The following day, SIS pieces were incubated with 0.167 mg/mL DNAse I (cat # 10104159001, Roche, Basel, Switzerland) in PBS with Ca^2+^ and Mg^2+^ (PBS^+^; cat # D8662, Sigma Aldrich, St. Louis, USA) for 2 hours at 37 °C under continuous agitation, then washed 3 times during 1 hour with PBS^—^. Samples for histopathological analysis, DNA quantification, and proteomics were collected as described above. SIS pieces were stored in PBS^—^ containing 1% P/S at 4 °C. Generated SIS scaffolds were γ-sterilized with a dosage of 25 kGy (Gammatom srl, Guanzate, Italy) and stored in PBS^—^ with 1% P/S at 4 °C until further use.

### 2.3 Generation of PerMa from porcine and human peritoneum

Peritoneal matrix (PerMa) was generated by decellularization of parietal peritoneum from the anterior abdominal walls from three different sources: pigs, autopsy of women with healthy peritoneum, and women with ovarian cancer undergoing primary cytoreductive surgery. Before the decellularization, samples for DNA quantification, proteomic analysis, and histopathological analysis were collected, as described for SIS. After sampling, the peritoneum was washed for a minimum of 1 hour and up to 18 hours with PBS^—^ with 1% P/S. Submesothelial fat was removed by blunt dissection to decrease the thickness of the membranes and improve decellularization efficiency. For decellularization, the peritoneum was incubated in 3.4% DOC dissolved in MQ water for 4 hours at 4 °C on a tilting platform. After decellularization, the peritoneum was washed in PBS^—^5 times for 1 hour at 4 °C, followed by treatment with 0.167 mg/mL DNAse I dissolved in PBS^+^ for 2 hours at 37 °C under continuous agitation. The peritoneum was then washed with PBS^—^3 times for 1 hour at 4 °C on a tilting platform. Decellularized tissue samples for histopathological analysis, DNA quantification, and proteomics were collected as described above. The remaining decellularized scaffold was stored in PBS^—^ with 1% P/S at 4 °C. Generated PerMa scaffolds were γ-sterilized with a dosage of 25 kGy (Gammatom srl, Guanzate, Italy) and stored in PBS^—^ with 1% P/S at 4 °C until further use.

### 2.4 Histology and immunohistochemistry

Specimens from native organs, decellularized scaffolds, and scaffolds repopulated with cells were fixed with 4% PFA for 1 hour at room temperature and embedded in paraffin. Formalin-fixed paraffin-embedded (FFPE) samples were cut into 3 µm thick sections.

#### Hematoxylin and eosin staining

Slides were deparaffinized in xylene for 20 minutes, rehydrated through 100%, 95%, and 70% ethanol baths (10 dips in each), and rinsed in water. Slides were incubated for 7 minutes in hematoxylin, then rinsed in water, dipped 8 times in 0.25% ammonia solution, rinsed for 5 minutes in water, incubated for 1 minute in eosin, rinsed by 10 dips in water, and immediately dehydrated through 70%, 95%, 100% ethanol baths, and xylene before mounting.

#### Elastica/Van Gieson

Deparaffinized and rehydrated samples were stained with an Elastica Van Gieson staining kit (Roche Tissue Diagnostics, Tucson, AZ, USA): 4 minutes incubation in hematoxylin, followed by 12 minutes incubation in FeCl Oxidizer & Lugol Iodine, washing with tap water, 8 minutes incubation in Differentiator, washing with tap water, and finally 4 minutes incubation in Van Gieson CS, dehydration and mounting.

#### Alcian Blue

Deparaffinized and rehydrated samples were stained with an Alcian Blue Staining Kit (Roche Tissue Diagnostics, Tucson, AZ, USA): 8 minutes incubation in Alcian Blue, washing, 4 minutes incubation in Alcian Blue nuclear fast red (NFR), and finally washing, dehydration and mounting.

#### Immunohistochemistry (IHC)

Deparaffinization, antigen retrieval, and staining were done with a Dako PT-link, following the Dako Flex+ protocol (Agilent Technologies, Santa Clara, CA, USA). Antigen retrieval was done with low or high pH, as detailed in Supplementary Table 1. Immunohistochemical staining was done with the antibodies listed in Supplementary Table 1. Sections were counterstained with hematoxylin.

### 2.5 DNA quantification

Specimens of native organs and decellularized scaffolds were lyophilized overnight, weighed and chopped into 1×1 mm pieces, and digested with Proteinase K (Qiagen, Hilden, Germany) for 3 hours at 56 °C. DNA was extracted using the DNeasy blood and tissue kit (cat# 69504, Qiagen, Hilden, Germany) according to manufacturer’s instructions. DNA content was quantified with the Quant-iT^TM^ PicoGreen^TM^ dsDNA Assay Kit (ThermoFisher Scientific, Waltham, MA, USA) according to the manufacturer’s instructions. Measurements were performed at an excitation of 480 nm and an emission of 520 nm using a SpectraMax Gemini EM plate reader (Molecular Devices, San Jose, CA, USA). DNA values were determined from a Lambda-DNA standard curve.

### 2.6 3D printed crowns

Culture plate inserts, referred to as crowns, and staining rings, were custom-designed and 3D printed with a Form 2 3D printer (Formlabs, Somerville, MA, USA), using High Temp resin (Formlabs, Somerville, USA). The dimensions are given in Supplementary Fig. 2, and stl-files are given as Supplementary files. The post-processing included 15 minutes of soaking in isopropyl alcohol, followed by drying with compressed air to remove excess print material and curing in a UV chamber in two rounds of 20 minutes. The crowns and staining rings were autoclaved before use for cell cultures.

### 2.7 Multiphoton microscopy

#### Sample preparation

Native organs were imaged on the day of sample collection. Decellularized scaffolds were imaged within one week after decellularization. For imaging, scaffolds were mounted on 3D printed crowns and placed in PBS or cell culture medium (Figure 3).

#### Imaging

For each tissue type, samples from three to five donors were imaged. Three random fields for each sample were imaged with an Olympus FluoView FVMPE-RS> multiphoton laser scanning microscope equipped with an Ultrafast Ti:Sapphire Laser (Olympus, Tokyo, Japan). Images were captured in water immersion with a 25x objective at 920 nm excitation and 515-560 nm for intrinsic two-photon excited fluorescence (TPEF) and 490-515 nm emission for second harmonic generation (SHG). Detector voltage was 400 V for TPEF and 450 V for SHG. The image field was 509×509 μm with a resolution of 1024×1024 pixels and a Z step size of 2 μm.

#### Image analysis

Image processing was performed in FIJI (National Institute of Health, Maryland, USA). Prior to image analysis, the images were cropped to 250×250 μm in four consecutive Z stacks of 10 μm depth starting at the surface of the tissue. Maximum projection images were generated from the Z stacks prior to further image analysis. Extracellular matrix patterns were analyzed using the ImageJ plugin TWOMBLI (The Workflow Of Matrix BioLogy Informatics v.1, github.com/wershofe/TWOMBLI) (33). The TWOMBLI parameters were: contrast saturation = 0.35, min line width = 1, max line width = 40, min curvature window = 5, max curvature window = 95, min branch length = 1, max display high-density matrix (HDM) = 255, and min gap diameter = 10. For each sample, the values for the three image positions were averaged.

### 2.8 Diffusion assay

Diffusion assay was performed as previously described (34). In short, FITC-dextran with an average molecular weight of 4 kD or 40 kD (cat # 46944 and FD40, Sigma Aldrich, St. Louis, MO, USA) was dissolved in PBS^—^ at a concentration of 10 µM, and low-molecular-weight fragments were removed with Amicon Ultra-4 Centrifugal Filters with a cut-off of 3 kD or 30 kD (cat# UFC8003 and UFC8030, Merck Millipore, Burlington, VT, USA). Scaffolds were mounted on 3D printed crowns to generate a Transwell-like system and incubated with PBS^—^ at 4 °C overnight. At the start of the experiment, 2200 µL fresh PBS^—^ was added to the outer compartment, and 1000 µL of 10 µM FITC-dextran solution was added to the inner chamber. The plate was placed in an incubator at 37 °C, 5% CO2 for the remainder of the experiment. At given time points (0, 5, 10, 20, 40, 60, 90, 120, 180, 240, 300, 360, 1440 minutes) 50 µL samples were taken from the outer compartment and the initial volume in the outer compartment was restored by addition of fresh PBS^—^. The fluorescence signal was measured at an excitation of 490 nm and an emission of 525 nm using a SpectraMax Gemini EM plate reader (Molecular Devices, San Jose, CA, USA). The amount of FITC-dextran that diffused across the scaffold over time was determined from a FITC-dextran standard curve. The apparent permeability coefficient (Papp), giving the passive permeability of the substance per area over time, was calculated as described before (35). Briefly, Papp is calculated using the following formula:

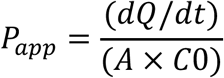

where dQ/dt is the rate of permeability (μmol/s), C0 is the initial concentration in the donor compartment, and A is the surface area of the filter (1.33 cm^2^).

### 2.9 Rheology

Rheological measurements were conducted using an Anton Paar Physica MCR302 (Anton Paar, Ostfildern, Germany) in a plate to plate set up equipped with a 25 mm diameter measuring probe, a solvent trap and a Peltier element. Samples were trimmed to fit the probe diameter and soaked with PBS^—^. The gap width was determined by lowering the probe until a standard force of 1 N was reached. Measurements were performed at 37 °C. Initially, a frequency sweep from 0.1 to 100 rad/s was performed at 0.5% strain deformation, followed by an amplitude sweep with increasing strain deformation from 0.01 – 500% at an angular frequency of 10 rad/s. Mean values of storage and loss modulus as well as the damping factor were calculated from the linear region of amplitude sweep measurements. The flow point was determined as shear stress at the intersection point of storage and loss modulus (G’ = G’’).

### 2.10 Proteomics

#### Cell lysis and protein digestion

10 mg of lyophilized native tissues and dECM scaffolds were weighed and chopped into 1×1 mm pieces. The samples were lysed in lysis buffer (100 mM Tris-HCl, pH 7.6, 5% SDS in MilliQ water) and sonicated 3-6 cycles of 30 seconds with 30 seconds rest on ice at power 30% (Sonics VibraCell CP130 sonicator, Cole-Parmer Instruments, Vernon Hills, IL, USA). The samples were boiled for 7 minutes at 95 °C and centrifuged for 10 minutes at 14000 x g. Protein lysates were transferred to new tubes. Protein concentration was measured using Pierce^TM^ BCA Protein Assay Kit (cat# 23227, ThermoFisher Scientific, Waltham, MA, USA) according to the manufacturer’s instructions.

#### On-beads precipitation and protein digestion

10 µg of the samples were precipitated with 70% acetonitrile onto magnetic beads (MagReSynAmine, Resyn Biosciences, Gauteng, South Africa). The proteins were washed on the beads with 100% acetonitrile, and 70% ethanol and then resuspended in 100 µL 50 mM ammonium bicarbonate. The proteins were reduced by the addition of 0.5 M dithiothreitol (DTT) to a final concentration of 10 mM and incubation at 56 °C for 30 minutes. Then, to alkylate proteins, 2.7 µL of 550 mM iodoacetamide (IAA) was added to a final concentration of 15 mM, and samples were incubated at room temperature in the dark for 30 minutes. 0.5 µg trypsin (Promega, Madison, WI, USA) was added to each sample for overnight on-beads protein digestion at 37 °C. The resulting peptides were concentrated and desalted before mass spectrometry analysis using the STAGE-TIP method with C18 resin disk (Affinisep).

#### LC-MS/MS analysis

Samples were analyzed by a nanoElute UHPLC coupled to a timsTOFfleX mass spectrometer (Bruker Daltonics, Bremen, Germany) via a CaptiveSpray ion source. Peptides were separated on a 25 cm reversed-phase C18 column (1.6 µm bead size, 120 Å pore size, 75 µm inner diameter, Ion Optics) with a flow rate of 0.3 µL/min and a solvent gradient from 0-35% B in 60 minutes. Solvent B was 100% acetonitrile in 0.1% formic acid and solvent A 0.1% formic acid in water. The mass spectrometer was operated in data-dependent Parallel Accumulation-Serial Fragmentation (PASEF) mode. Mass spectra for MS and MS/MS scans were recorded between m/z 100 and 1700. Ion mobility resolution was set to 0.85–1.30 V·s/cm over a ramp time of 100 ms. Data-dependent acquisition was performed using 4 PASEF MS/MS scans per cycle with a near 100% duty cycle. A polygon filter was applied in the m/z and ion mobility space to exclude low m/z, singly charged ions from PASEF precursor selection. An active exclusion time of 0.4 min was applied to precursors that reached 20,000 intensity units. Collisional energy was ramped stepwise as a function of ion mobility.

#### LC-MS/MS data analysis

MS raw files were submitted to MaxQuant software version 2.4.7.0 (36) for protein identification and label-free quantification. Parameters were set as follow: fixed modification: carbamidomethylation (C), protein N-acetylation and methionine oxidation as variable modifications. First search error window of 20 ppm and main search error of 4.5 ppm. Trypsin without proline restriction enzyme option was used, with two allowed miscleavages. Minimal unique peptides were set to 1, and false discovery rate (FDR) allowed was 0.01 (1%) for peptide and protein identification. For porcine samples the sus scrofa trembl and Swissprot (downloaded 2023) databases were used and for human samples the homo sapiens Swissprot database from uniprot (downloaded 2022) was used. The function match-between-runs were deactivated. Generation of reversed sequences were selected to assign FDR rates.

For statistical comparison analysis, Perseus version 2.0.11 was used. Intensities were log2 transformed and data was filtered based on 50% valid values in at least one group, missing values were imputed from normal distribution and student t-test was performed with permutation-based FDR cutoff of p-value<0.05.

#### Matrisome matching

Protein ID from MaxQuant proteinGroup file was mapped to gene names using the uniprot protein ID mapping. Around 40% of the protein IDs could not be mapped. The gene names were matched to the matrisome database (37).

#### Data availability

The mass spectrometry proteomics data have been deposited to the ProteomeXchange Consortium via the PRIDE (38) partner repository with the dataset identifier PXD049045.

### 2.11 SIS and PerMa cell cultures

#### Cell culture

Scaffolds were mounted as a Transwell-like system (Fig. 3C) and placed in 12 well plates in cell culture medium at 37 °C, 5% CO2 overnight. The following day, 1.5 mL medium was added to the outer chamber, and cells were seeded in the inner chamber in a volume of 1 mL. For SIS monocultures, ovarian cancer cells were seeded on the submucosal surface. For PerMa, cells were seeded on the layer facing the peritoneal cavity. The cells were cultured at 37 °C, 5% CO2. All EOC cell lines were GFP+ and the BJ fibroblasts were miRFP670+. The cell culture experiments were performed two times in triplicates, unless otherwise stated.

#### Imaging

Scaffolds were imaged on days 3, 7, 10, 14 and 21 using a Dragonfly 505 confocal spinning disk system (Andor Technologies, Inc, Belfast, Northern Ireland). To enable imaging, scaffolds were flipped so that the cells were on the outer surface of the scaffold on day 3 and kept in that position for the remaining period of the experiment. For each scaffold, three representative imaging fields were selected and imaged at 10x magnification, with a Z-stack depth of 150 to 300 µm and a 2 µm step size. The background matrix structure was imaged using a 561 nm laser line and a 575-625 nm filter. GFP+ cells were imaged using a 488 nm laser line and a 500-550 nm filter. For cocultures on PerMa, BJ miRFP670+ fibroblasts were added to the culture and imaged using a 637 nm laser line and a 663-738 nm filter.

#### Image analysis

Image processing was performed in IMARIS software v. 9.9 (Bitplane, Zurich, Switzerland). Quantification of fluorescent signal was analyzed using the surfaces function.

### 2.12 Drug screen and viability assay

Carboplatin (cat # 507662, 10 mg/mL, Fresenius Kabi, Bad Homburg, Germany) was used within 48 hours after the seal was broken. Drug dilution series were prepared using cell culture medium. OV90 cells were used for all drug sensitivity assays.

To determine carboplatin EC50 values for the OV90 cell line cultured in a 2D monolayer, WST-1 assay was performed (cat # 11644807001, Roche, Basel, Switzerland). 2000 cells/well were seeded in 96 well cell culture plates and allowed to attach for 24 hours. The following day, the culture medium was removed, and carboplatin at different concentrations was added onto the plate. After 72 hours, WST-1 reagent was added (1:10) to each well and incubated for 2 hours at 37 °C, 5% CO2. The absorbance of formazan was measured at 440 nm with a Spectra Max Plus 384 microplate reader (Molecular Devices, San Jose, USA). The experiment was performed in triplicate and repeated three times.

Carboplatin sensitivity for OV90 in 2D culture was also assessed using apoptosis and necrosis as markers for cell viability. Five doses (10 µM, 31.6 µM, 45.6 µM (= EC50), 100 µM, and 316 µM) were chosen based on the WST-1 assay results. 90,000 cells/well were seeded in 6 well culture plates in 2 mL volume and allowed to attach for 24 hours. The following day, the culture medium was removed, and Carboplatin at different concentrations was added onto the plate at a volume of 3 mL per well. After 72 hours, the medium from each well was collected in individual tubes. The remaining attached cells were then trypsinized and combined with the medium containing detached cells. The cells were stained with Annexin V/PI (cat # V13245, Invitrogen, Waltham, MA, USA) and analyzed by flow cytometry for detection of apoptotic and necrotic cells. The experiment was performed with three technical replicates for each drug dose.

Based on the EC50 value for OV90 and the results from the apoptosis/necrosis assay, three drug doses (10 µM, 50 µM, and 100 µM) were chosen for further drug sensitivity testing in 3D culture. PerMa was mounted as a Transwell-like system and placed in 12 well plates in cell culture medium at 37 °C, 5% CO2 overnight. The following day, 25,000 cells/scaffold were seeded. On day three after seeding, the scaffolds were flipped, leaving the cells on the outer surface. On day seven after seeding, the PerMa scaffolds were imaged using a Dragonfly 505 confocal spinning disk system (Andor Technologies). Following imaging, the scaffolds were moved to a new 12-well plate containing carboplatin at different doses and cultured at 37 °C, 5% CO2. On day ten after seeding, the PerMa scaffolds were stained with an apoptosis/necrosis assay kit (cat # ab176749, Abcam, Cambridge, United Kingdom). Staining was performed according to the manufacturer’s recommendations using the 3D-printed staining ring. Briefly, the staining reagents and washing buffers were added in a volume of 300 µL to the staining ring, and the crowns with PerMa were placed on top of the staining ring. Staining was performed according to manufacturer’s recommendations. Scaffolds were washed in washing buffer for 1 minute, stained with 7-AAD, Apopxin Green, and CytoCalcein Violet for 30 minutes, followed by 3 x 1 minute washing in washing buffer. The scaffolds were imaged using the Dragonfly confocal microscope. Images were taken at 10x magnification, with a Z-stack depth of 150 to 300 µm with a 2 µm step size. Three representative imaging fields were selected for each scaffold. 7-AAD was imaged using a 561 nm laser line and a 575-625 nm filter. Apopxin Green was imaged using a 488 nm laser line and a 500-550 nm filter. CytoCalcein Violet was imaged using a 405 nm laser line and a 425-475 nm filter. Image analysis was performed in the IMARIS image analysis software using the surfaces and spots functions.

### 2.13 CAR T cytotoxicity testing

TAG72 CAR single chain variable fragment (scFv), huCC49, was derived from Kashmiri et al. (39). Muc16 CAR scFv (K101) was generated as described in Casey et al. (40). Both CARs are second-generation containing a CD8 hinge and CD8 transmembrane domain, a 4-1BB intracellular domain, and CD3ζ (41). Retroviral particles carrying the CAR vectors and transduction of T cells were done as described previously (42). Briefly, HEK cells were transfected with packaging vectors and a retroviral vector encoding the CAR constructs. After 2 days of harvesting, the supernatant, containing viral particles, was used in two rounds of transduction of PBMCs activated for 48 hours with anti-CD3 and anti-CD28 antibodies (coated on a 24-well plate at 1 µg/mL respectively). CAR expression on the T cells was assessed with an anti-G4S linker (Cell Signaling Technology, US) 3 and 8 days after transduction.

Cell cultures were prepared as described in section 2.11. Briefly, 25,000 OV90 cells were seeded per scaffold and treated with CAR T cells targeting TAG-72 or Mock T cells. The scaffolds were flipped and imaged with a Dragonfly 505 confocal microscope after three and seven days to confirm cell growth. On day eight, the scaffolds containing OV90 cells were flipped back so the cells were again on the inner surface and placed in new wells containing 1.5 mL medium. CAR T or mock T cells were labeled with CellTrace^TM^ Violet Cell Proliferation Kit (CTV; cat # C34557, Invitrogen, Waltham, USA) according to the manufactureŕs protocol and seeded in a volume of 0.5 mL to the inner chamber. The scaffolds were incubated at 37 °C, 5% CO2. After 24 hours, the scaffolds were flipped, so the cells were again on the outer surface and imaged with a Dragonfly 505 confocal microscope, as described above. CAR T/mock T cells were imaged using a 405 nm laser line and a 425-475 nm filter and GFP positive EOC cell lines using a 488 nm laser line and a 500-550 nm filter. Image analysis was performed in the IMARIS image analysis software using the surfaces function.

### 2.14 Statistical analyses

Statistical analyses were performed in PRISM Graphpad (version 10.1.0). Data is presented as mean ± standard deviation. For analyses comparing two datasets, unpaired t test was used. For comparison of more than two datasets, analysis of variance (ANOVA) with Tukey’s multiple comparison test was used. Significance was set to p < 0.05. Details about statistical analyses, including p values, are presented in the figure legends.

## 3 Results

### 3.1 Chemical and enzymatic decellularization of peritoneum results in a cell-free ECM scaffold

3D *in vitro* organotypic models can be generated by culturing normal and malignant cells on decellularized tissue scaffolds, such as the porcine small intestinal submucosa (SIS) scaffold (30, 31). To establish a model that mimics peritoneal HGSOC metastases, we developed a decellularized tissue scaffold from porcine peritoneum, referred to as pPerMa. We compared this to the previously established porcine SIS (pSIS) and decellularized human peritoneum obtained from either autopsy of patients with no intra-abdominal disease (hPerMa) or surgery of EOC patients (ocPerMa; Fig. 1a).

**Figure 1.**
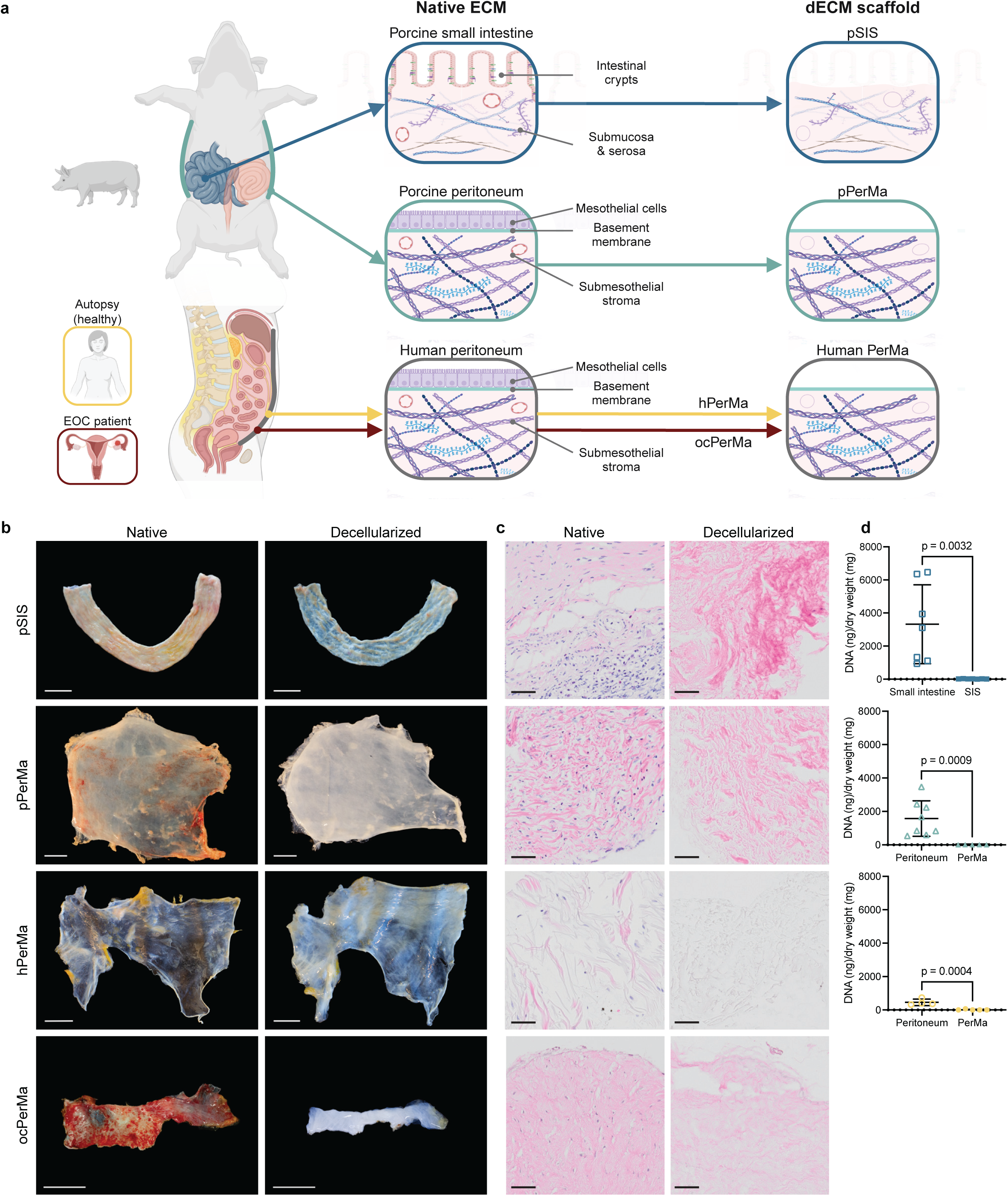
Decellularization of small intestine and peritoneum generates dECM scaffolds. **a** Schematic illustration of the generation of dECM scaffolds from porcine small intestine and peritoneum, and human peritoneum. **b** Photographs of porcine small intestine and porcine and human peritoneum before (left) and after (right) decellularization. Scale bars 30 mm. **c** Corresponding hematoxylin and eosin (H&E) staining of native and decellularized tissues. Scale bars 50 µm. **d** DNA content of native and decellularized porcine small intestine (n = 7 native and n = 7 decellularized), porcine peritoneum (n = 8 native and n = 8 decellularized), and human peritoneum (n = 4 native and n = 6 decellularized). Data represent mean ± SD, unpaired two-tailed t-test.

Porcine and human parietal peritoneum were collected from the anterior abdominal walls and decellularized using a chemical detergent followed by enzymatic DNA degradation (Supplementary Figure 1). We chose the detergent sodium deoxycholate (SDC) since it has been shown to effectively decellularize tissues of various thicknesses without compromising the structure of the ECM (43–45). The SIS scaffold was prepared as described previously (32).

To evaluate the decellularization efficacy, we assessed cell removal by histology and residual DNA content. Samples were taken from native intestine and peritoneum as well as from small intestinal submucosa (SIS) and peritoneal matrix (PerMa; Fig. 1b). The decellularization resulted in a cell-free scaffold as demonstrated by hematoxylin and eosin (H&E) staining (Fig. 1c). DNA quantification confirmed depletion of cell nuclei and genetic material to an average 20.03 ± 10.97 ng/mg in pSIS, 2.91 ± 4.62 ng/mg in pPerMa, and 12.48 ± 26.95 ng/mg in hPerMa (Fig. 1d). Altogether, the decellularization protocol results in an efficient removal of cells and a significant decrease in DNA content.

### 3.2 A custom-designed culture plate insert enables characterization and cell culture

To enable the characterization and the establishment of cell cultures with the dECM scaffolds, we engineered a 3D printed culture plate insert, referred to as a crown with attachment clip (Fig. 2a and Supplementary Fig. 2a,b). The crown serves three key functions: 1) elevating and securing the dECM scaffold above the surface of a multiwell culture plate, 2) establishing a Transwell®-like system where two chambers are separated solely by the scaffold, and 3) enabling confocal imaging, ensuring direct contact between the scaffold and the imaging surface. To enable the staining of scaffolds in a lower volume, we developed a 3D-printed staining ring (Fig. 2b, Supplementary Fig. 2c). The crowns and staining ring were developed to fit 12-well plates.

**Figure 2.**
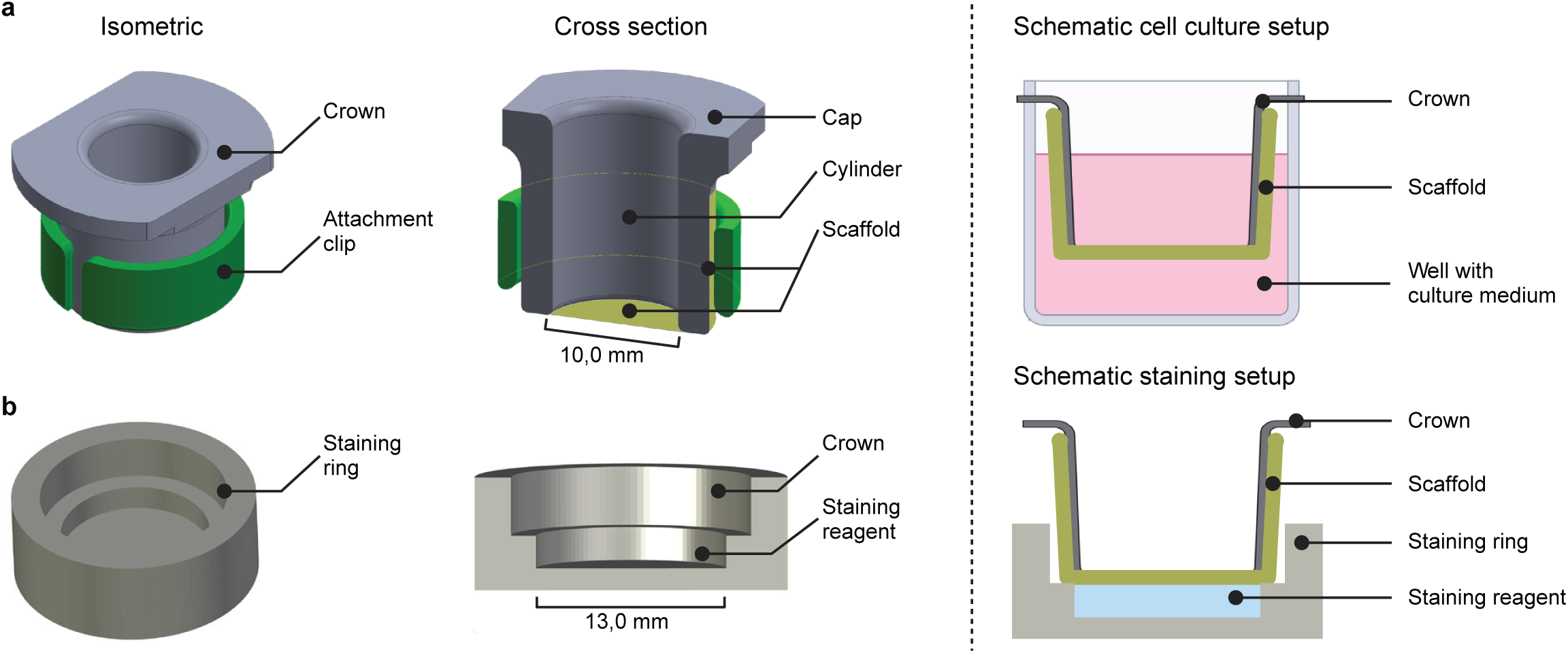
Custom-designed 3D-printed culture plate inserts and staining rings enable cell culture, staining, and confocal imaging of 3D cultures using dECM scaffolds. **a** A 3D-printed culture plate insert, referred to as a crown, suspends and secures the dECM scaffold above the surface of a culture plate well, creating a Transwell®-like setup. **b** A staining ring enables staining of the dECM scaffold in a volume of 300 µL.

**Figure 3.**
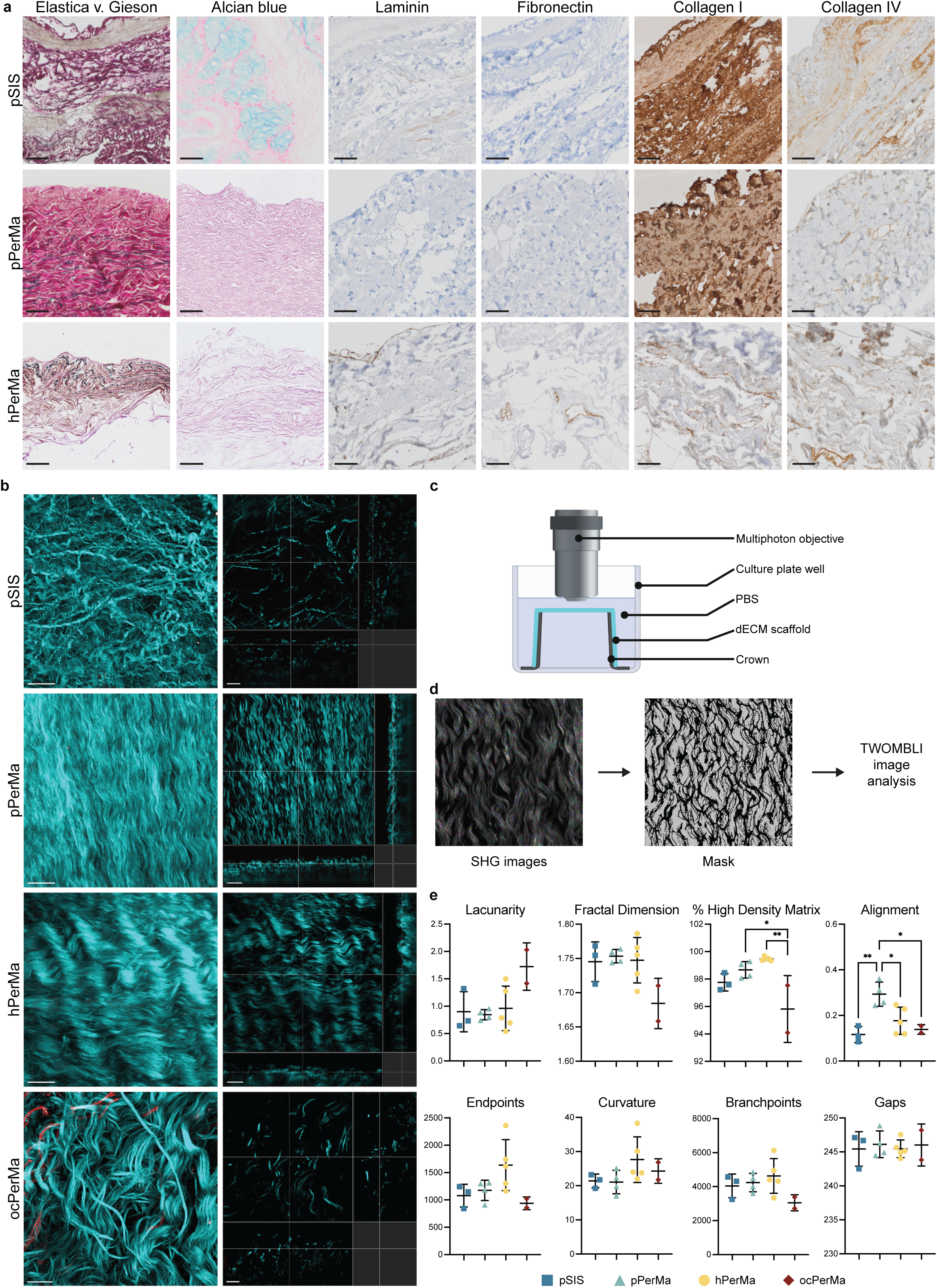
The structure of major ECM proteins differs between the dECM scaffolds. **a** Representative images of Elastica v. Gieson and Alcian blue staining and immunohistochemical staining of laminin, fibronectin, collagen I, and collagen IV in pSIS, pPerMa, and hPerMa. Scale bars 50 µm. **b** Representative second harmonic generation (SHG) microscopy images of pSIS, pPerMa, hPerMa, and ocPerMa. 3D renderings (left) and single optical sections (right). Scale bars = 50 µm. **c** Schematic illustration of the setup for multiphoton imaging. The dECM scaffolds are mounted on crowns and placed bottom-up in a 6-well plate filled with PBS. The multiphoton objective is immersed in PBS. **d** Example images showing the masking of SHG images for The Workflow Of Matrix BioLogy Informatics (TWOMBLI) image analysis. **e** TWOMBLI analysis results. Data represent mean ± SD. *P < 0.05, **P < 0.01, one-way ANOVA with Tukey’s multiple comparison test. N = 3 for pSIS, n = 4 for pPerMa, n = 5 for hPerMa, n = 2 for ocPerMa. Three representative images were captured and analyzed for each sample. Images were cropped to 250 µm (x) * 250 µm (y) * 10 µm (z) stacks. Four z stacks were generated per image area to include the top 40 µm from the surface.

#### Structure Design and Dimensions

Our design centered on a cylindrical structure, optimizing gas and fluid access to the dECM scaffold using conventional pipetting techniques. The inner chamber has a diameter of 10 mm, complemented by a wall thickness of 2.5 mm (Fig. 2a, Supplementary Fig. 2a). This thickness was strategically chosen based on the material’s properties, balancing the ease of printing and processing with durability for repeated use.

#### Well Plate Compatibility and Usability Features

To prevent the dECM scaffold from contacting the bottom of the well, we incorporated a cap with a larger diameter (24 mm), exceeding the well diameter of the 12-well plate (Fig. 2a, Supplementary Fig. 2a). This design prevents interference with adjacent wells.

#### Centering and Depth Considerations

To ensure the crown’s centered positioning within the well, we added a notch in the cap at a diameter of 20.5 mm (0.5 mm smaller than the well’s diameter; Supplementary Fig. 2a). The main cylinder’s depth was set at 13.00 mm to avoid contact with the well bottom. Where feasible, we added fillets of 1 mm or 0.5 mm to mitigate any potential damage to the dECM scaffold during application.

#### Retention Mechanism

A critical aspect of the design was a small “attachment clip,” functioning similarly to an open C-clip (Fig. 2a, Supplementary Fig. 2b). This clip leverages the flexibility of the 3D printed material to exert sufficient clamping force on the dECM scaffold, ensuring secure retention while allowing for easy removal without damage to the dECM scaffold. The clip’s wall thickness is 1.5 mm, fine-tuned for optimal clamping force. To further protect the dECM scaffolds, the retaining clip includes fillets of 0.5 and 1.0 mm to prevent puncturing during application.

#### Staining Ring

The staining ring has a chamber with a volume of 300 µL that is filled with staining solution. The crown is placed on top of this chamber, supported on the sides.

FormLabs High Temp Resin (FLHTAM01) can withstand autoclave temperatures (46) and has previously been demonstrated to be biocompatible after post-print treatment (47). We did not observe adverse effects of the material on cell viability.

### 3.3 The dECM scaffolds retain the structure of the native tissues and show similarities in structure

To assess the integrity of the ECM after tissue decellularization, we compared histological staining of major ECM components and the structure of the ECM with multiphoton microscopy (Fig. 3 and Supplementary Fig. 3). Histological staining showed similar patterns of laminin, fibronectin, collagen I, and collagen IV positivity in the native (Supplementary Fig. 3a) and decellularized (Fig. 3a) tissues from the same source. Second harmonic generation (SHG) and two-photon excitation fluorescence (TPEF) by multiphoton microscopy allow label-free visualization of non-centrosymmetric structures (typically collagen fibers) and tissue autofluorescence, respectively (48) (Fig. 3b). For multiphoton imaging, the scaffolds were mounted on crowns, placed in 6 well plates with the tissue facing up, and submerged in PBS (Fig. 3c). To assess the effect of decellularization on the matrix patterns, we used the ImageJ plugin TWOMBLI to quantitatively analyze the multiphoton images of our samples (33) (Fig. 3d). Fiber lacunarity, fractal dimension, % high-density matrix (HDM), branchpoints, and endpoints all remained unaltered between native and decellularized samples of the same tissue type (Fig. 3e). Only matrix alignment increased in the decellularized pPerMa (0.2931 ± 0.0531) compared to the native porcine peritoneum (0.1109 ± 0.0492), p = 0.0003 and curvature decreased in the decellularized pSIS (21.41 ± 2.011) compared to the native porcine intestine (25.44 ± 0.9989), p = 0.0358 (Supplementary Fig. 3c). In summary, the decellularization preserved the structure of major ECM components.

We also used histological staining and multiphoton microscopy to assess the differences between dECM scaffolds derived from different tissues. Alcian Blue stained positive only in pSIS and not pPerMa or hPerMa (Fig. 3a). Elastica/Van Gieson stain demonstrated elastic fibers interspersed between collagen fibers in the submesothelial stroma of human and porcine PerMa. The elastic fibers of pSIS were located deeper in the tissue. Immunohistochemical staining showed positivity for laminin, fibronectin, collagen I, and collagen IV in hPerMa (Fig. 3a). Collagen I and collagen IV stained positive in pSIS and pPerMa, while laminin stained only weakly positive in pSIS, and fibronectin did not stain in pSIS nor pPerMa. Of note, the content of high-density matrix, as assessed by TWOMBLI in the multiphoton images, was significantly lower in ocPerMa (95.82% ± 2.440) compared to hPerMa (99.47% ± 0.140, p = 0.0005) and pPerMa (98.67% ± 0.599, p = 0.0101) (Fig. 3e). Instead, the matrix had significantly more fiber alignment in pPerMa (0.2931 ± 0.0531) compared to pSIS (0.1163 ± 0.0353, p = 0.0047), hPerMa (0.1765 ± 0.0595, p = 0.0273) and ocPerMa (0.1388 ± 0.0211, p = 0.0238) (Fig. 3e). No significant differences were observed between the dECM scaffolds in lacunarity, fractal dimension, curvature, branchpoints, or endpoints (Fig. 3e). Overall, the structure of major ECM components was most comparable in the dECM scaffolds from different normal tissue sources. Of note, the ocPerMa displayed a reduced high-density matrix compared to the others, however it should be noted the small number of just two tissues.

### 3.4 PerMa is a viscoelastic, permeable scaffold

Next to structure, the physical properties of the ECM, such as elasticity or porosity are known to influence cellular behaviors, including spreading, growth, proliferation, migration, and differentiation (49). Therefore, we investigated whether the scaffolds differ in their physical characteristics. Oscillatory rheological measurements were conducted to assess the mechanical properties of the pPerMa in comparison to the pSIS and the hPerMa.

Frequency sweep measurements revealed a similar response of all three scaffolds to increasing angular frequencies (Fig. 4b). Storage modulus (G’), which is a measure of the elastic properties of a material, and loss modulus (G’’), describing the viscous material portion, exhibited parallel runs, confirming a viscoelastic solid behavior of all scaffolds. Shear moduli G’ and G’’ differed among the scaffolds (hPerMa < pSIS < pPerMa), indicating different viscoelastic properties. Strain sweep measurements confirmed this presumption (Fig. 4c). The pPerMa exhibited a significantly higher storage and loss modulus (846 ± 100 Pa and 122 ± 26 Pa) compared to the pSIS (464 ± 167 Pa and 60 ± 31 Pa) and the hPerMa (207 ± 156 Pa and 41 ± 27 Pa), demonstrating that the pPerMa is more rigid than hPerMa and the pSIS (Fig. 4d,e). Next, the damping factor (tan δ = G’’ / G’) was calculated to assess the elastic and viscous proportions of the scaffolds. The pPerMa (0.14 ± 0.02) and the pSIS (0.12 ± 0.02) exhibited a significantly lower damping factor compared to the hPerMa (0.23 ± 0.08) (Fig. 4f), suggesting an increased portion of elastic components in the pPerMa. This was corroborated by analysis of the flow point, defining the transition from solid to viscous behavior (G’ = G’’) of a viscoelastic material. For both, pPerMa (53 ± 14 Pa) and pSIS (46 ± 15 Pa), a significantly higher shear stress (τ) was required to reach the flow point compared to the hPerMa (18 ± 15 Pa) (Fig. 4g), indicating that both, pPerMa and pSIS contain more elastic components than the hPerMa. Summarized, the pPerMa is more rigid than the pSIS and the hPerMa while the viscoelastic behavior is more similar to the porcine SIS.

**Figure 4.**
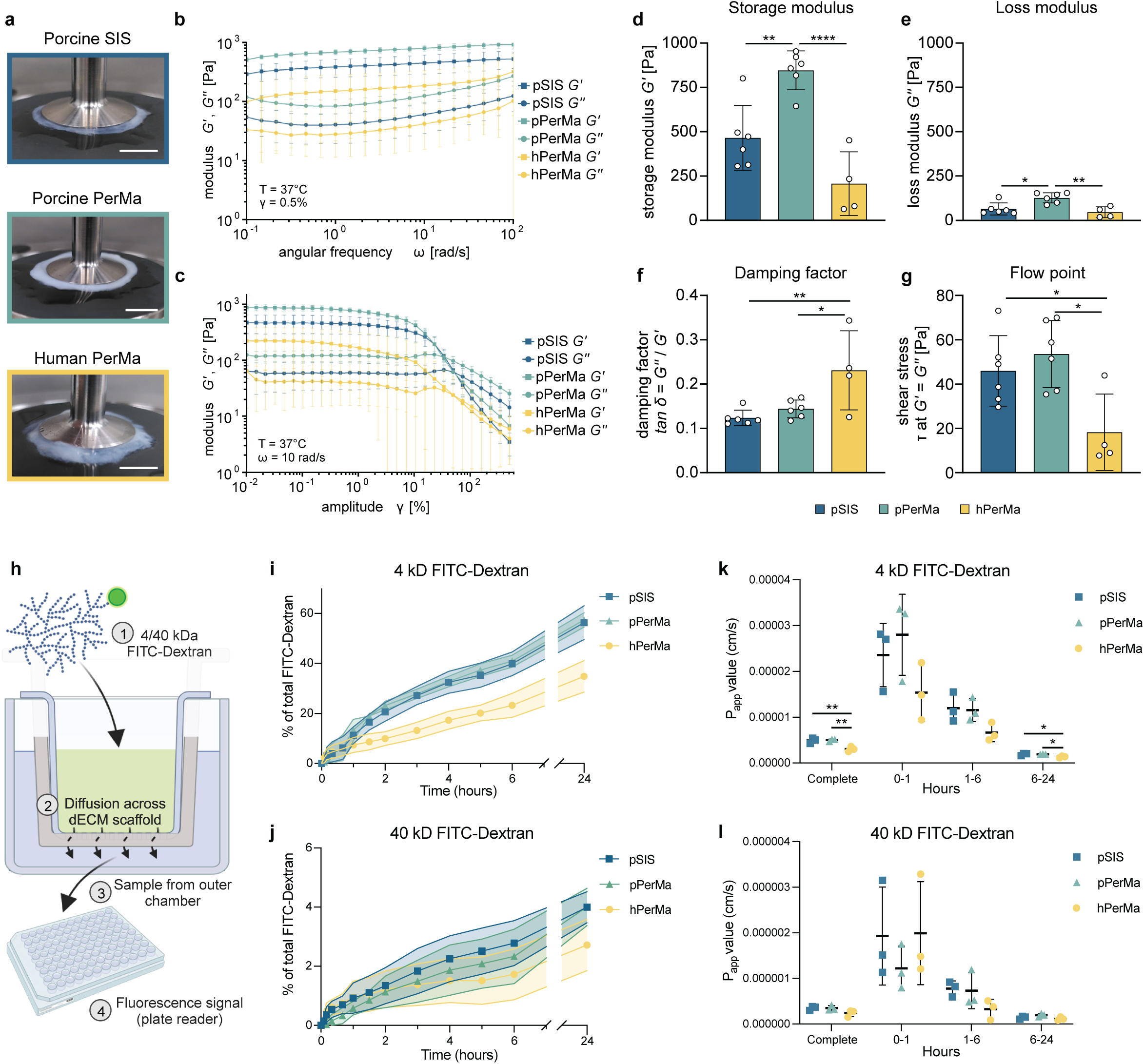
Rheological analyses reveal differences in the stiffness and permeability of the dECM scaffolds. **a** Photographs of porcine SIS (blue), porcine PerMa (green), and human PerMa (yellow) mounted in the plate-to-plate rheometer. **b,c** Data plot of oscillatory measurements showing the storage (G’, square) and loss (G’’, circle) modulus of the pSIS, the pPerMa, and the hPerMa. Moduli are shown as a function of frequency (b, γ = 0.5%) and strain deformation (c, ω = 10 rad/s). **d-g** Rheological parameters of the dECM scaffolds calculated from the linear region of the amplitude sweep. Single data points represent measurements of individual biological samples. Data represent mean ± SD. *P < 0.05, **P < 0.01, ****P < 0.0001, one-way ANOVA with Tukey’s multiple comparison test. **h** Schematic illustration of the diffusion assay. The dECM scaffolds were mounted in a Transwell®-like setup. 4 or 40 kD FITC-Dextran was added to the inner chamber, and samples were taken from the outer chamber at different time points (0, 5, 10, 20, 40, 60, 90, 120, 180, 240, 300, 3600, and 1440 minutes). The fluorescence signal was analyzed with a plate reader. **i,j** Diffusion of 4 kD (i) and 40 kD (j) FITC-Dextran across the scaffolds over time. Data represent mean ± SD displayed as error shadows. The experiment was performed three times with three replicates for each scaffold. **k,l** The apparent permeability coefficient (Papp) for 4 kD (j) and 40 kD (k) FITC-Dextran calculated for different time frames. *P < 0.05, **P < 0.01, one-way ANOVA with Tukey’s multiple comparison test.

To evaluate scaffold permeability, we analyzed the diffusion of molecules with low (4 kDa) and high (40 kDa) molecular weights across the scaffolds. Decellularized scaffolds were mounted in a Transwell-like setup with an inner and an outer chamber separated only by the scaffolds (Fig. 4h). 4 kDa or 40 kDa FITC-Dextran were added to the inner chamber, and samples were taken from the outer chamber at different time points. Diffusion of FITC-Dextran across the scaffold was calculated as a percentage of total FITC-Dextran. The volume of the outer chamber was 68.75% of the total volume. All three matrices were permeable to low molecular weight FITC-Dextran (Fig. 4i). At 24 hours, significantly less 4 kDa FITC-Dextran had diffused across hPerMa, compared to pSIS and pPerMa (34.83% vs. 56.33% and 57.45%, respectively). The diffusion rate, given by the apparent permeability coefficient (Papp), was significantly lower for the hPerMa compared to pSIS and pPerMa for the complete period (p = 0.0078 and p = 0.0061, respectively; Fig. 4k). The matrices were only partially permeable to high molecular weight FITC-Dextran (Fig. 4j). At 24 hours, 4.00% for pSIS, 4.01% for pPerMa, and 2.72% for hPerMa of 40 kDa FITC-Dextran diffused, with no statistically significant difference in amount diffused FITC-Dextran or Papp value. For both low and high molecular weight FITC-Dextran, the diffusion rate was highest within the initial 60 minutes (Fig. 4 k,l). Interestingly, diffusion of 4 kD across the human PerMa was slower compared to the pPerMa and pSIS, indicating a lower permeability of the hPerMa.

### 3.5 The dECM scaffolds have unique matrisome compositions

To characterize the biomolecular composition of the native tissues and decellularized scaffolds, we performed label-free quantitative proteomics by liquid chromatography-tandem mass spectrometry (LC-MS/MS). The number of identified proteins varied between the groups ranging from approximately 2000 to 7000 proteins and was consistently lower in the dECM scaffolds compared to the corresponding native tissues (Supplementary Fig. 4a). This is expected after the removal of cellular content (50). The number of identified proteins was significantly higher in pSIS compared to pPerMa, hPerMa, and ocPerMa (Supplementary Fig. 4b).

Principal component analysis (PCA) showed that the biological replicates were similar (Fig. 5a,b). Both native and decellularized replicates clustered into separate groups, showing the reproducibility of the decellularization method. While the native human peritoneum showed variation between replicates, the biological replicates of decellularized hPerMa were more similar.

**Figure 5.**
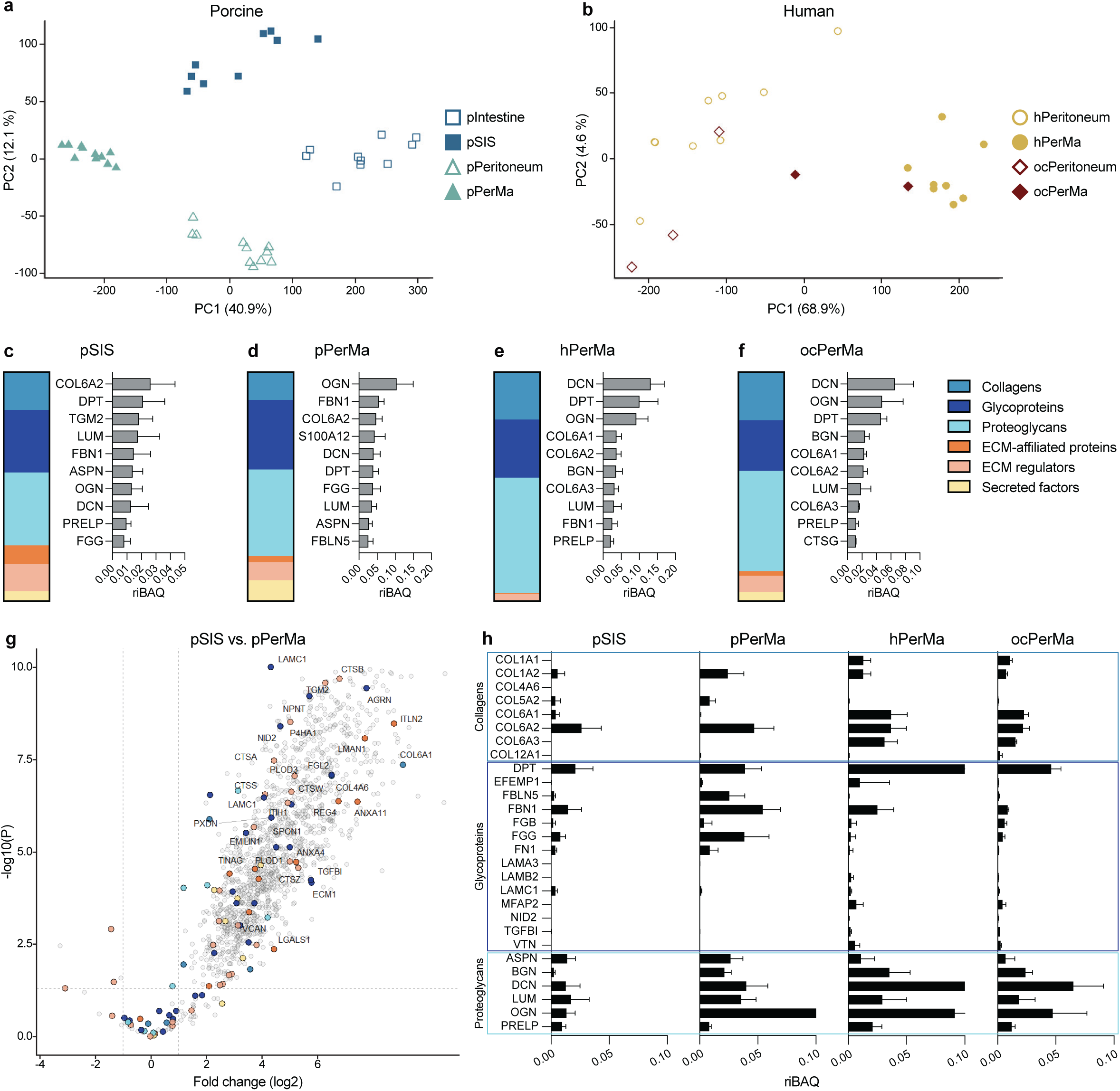
Proteomic analysis by liquid-chromatography mass spectrometry (LC-MS/MS). **a, b** Principal component analysis (PCA) plots of the total proteome for porcine (a) and human (b) samples. **c-f** Composition of total matrisome proteins and the ten most abundant matrisome proteins in pSIS (n = 10), pPerMa (n = 11), hPerMa (n = 8), and ocPerMa (n = 2). The ratios of the top ten proteins are given as iBAQ intensity relative to the total iBAQ intensity (riBAQ) for each sample. Data represent mean ± SD. **g** Volcano plot of protein intensities in pSIS (n = 10) vs. pPerMa (n = 11). Matrisome proteins with at least a 3-fold change (log2) are labeled. **h** Profile bar charts for pSIS (n = 10), pPerMa (n = 11), hPerMa (n = 8), and ocPerMa (n = 2) of selected core matrisome proteins (n = 28). The proteins are sorted into collagens (top box), glycoproteins (middle box), and proteoglycans (bottom box). The ratios are given as relative iBAQ intensities. Data represent mean ± SD.

To evaluate whether key ECM proteins were retained after decellularization, we compared the abundance of core matrisome proteins and matrisome-associated factors, as defined by Naba et al. (37), in native and decellularized samples. The relative abundance of matrisome and matrisome-associated proteins vs. non-matrisome proteins was higher in all dECM scaffolds compared to their native counterparts (Supplementary Fig. 4c). The proportion of core matrisome and matrisome-associated proteins increased from 12% in pIntestine to 18% in pSIS, from 27% in pPeritoneum to 57% in pPerMa, from 28% in hPeritoneum to 66% in hPerMa, and from 13% in ocPeritoneum to 45% in ocPerMa. Concordance was observed between native and decellularized tissues when comparing the 10 most abundant matrisome proteins (Fig. 5c-f, Supplementary Fig. 4d). Volcano plots comparing native and decellularized tissues revealed enrichment of particularly core matrisome proteins in the dECM scaffolds. However, there was also a depletion of several matrisome proteins, especially matrisome-associated proteins (Supplementary Fig. 4 e-g). In summary, the decellularization resulted in the enrichment of matrisome proteins and retained the most abundant ECM proteins.

We then compared the different dECM scaffolds. The composition of matrisome and matrisome-associated proteins was similar across tissues, with some organ-specific differences (Fig. 5c-f). All dECM scaffolds were rich in glycoproteins and proteoglycans. The highest proportion was seen in hPerMa, where glycoproteins and proteoglycans together constituted 76% of the total matrisome proteins (Fig. 5e). This proportion was 59% for pSIS, 68% for pPerMa, and 66% for ocPerMa. The samples displayed great diversity in the composition of different categories of matrisome proteins. Collagen Type VI (COL6A1-3), glycoproteins such as dermatopontin (DPT), and proteoglycans such as lumican (LUM) and decorin (DCN) were found among the top 10 most abundant matrisome proteins. Several of these were identified in the top 10 lists across all groups. Despite the similarities in matrisome composition across all dECM scaffolds, the volcano plot comparing pSIS and pPerMa revealed differences in the abundance of non-matrisome and matrisome proteins (Fig. 5g). A panel of 28 core matrisome proteins revealed further differences in the relative abundance between the dECM scaffolds (Fig. 5h).

In summary, porcine PerMa represents a biologically relevant tissue scaffold with similarities to human PerMa in structure, physical properties, and biomolecular composition.

### 3.6 The pPerMa and the pSIS enable 3D in vitro cell cultures

Porcine tissues have several advantages over human tissues, including availability and less batch-to-batch variation. Therefore, porcine scaffolds were used to establish three-dimensional (3D) in vitro cell cultures. We first investigated the cytocompatibility of pSIS and pPerMa, and tested next, whether the ECM-specific differences in mechanical properties and protein composition of pSIS and pPerMa affected cellular behavior *in vitro*.

The matrices were mounted on the crowns, and cells were seeded on the surface of the inner chamber (Fig. 6a). Three EOC cell lines – OV90, Caov-3, and COV318 – were chosen based on their genetic resemblance to HGSOC (51). After three days, the scaffolds were flipped, leaving the cells on the outer surface where they were accessible for confocal imaging (Fig. 6a). The scaffolds were imaged on days 3, 7, 10, 14, and 21 after seeding (Fig. 6a). The surfaces function in the IMARIS image analysis software was used to estimate cell volume based on the fluorescent signal.

**Figure 6.**
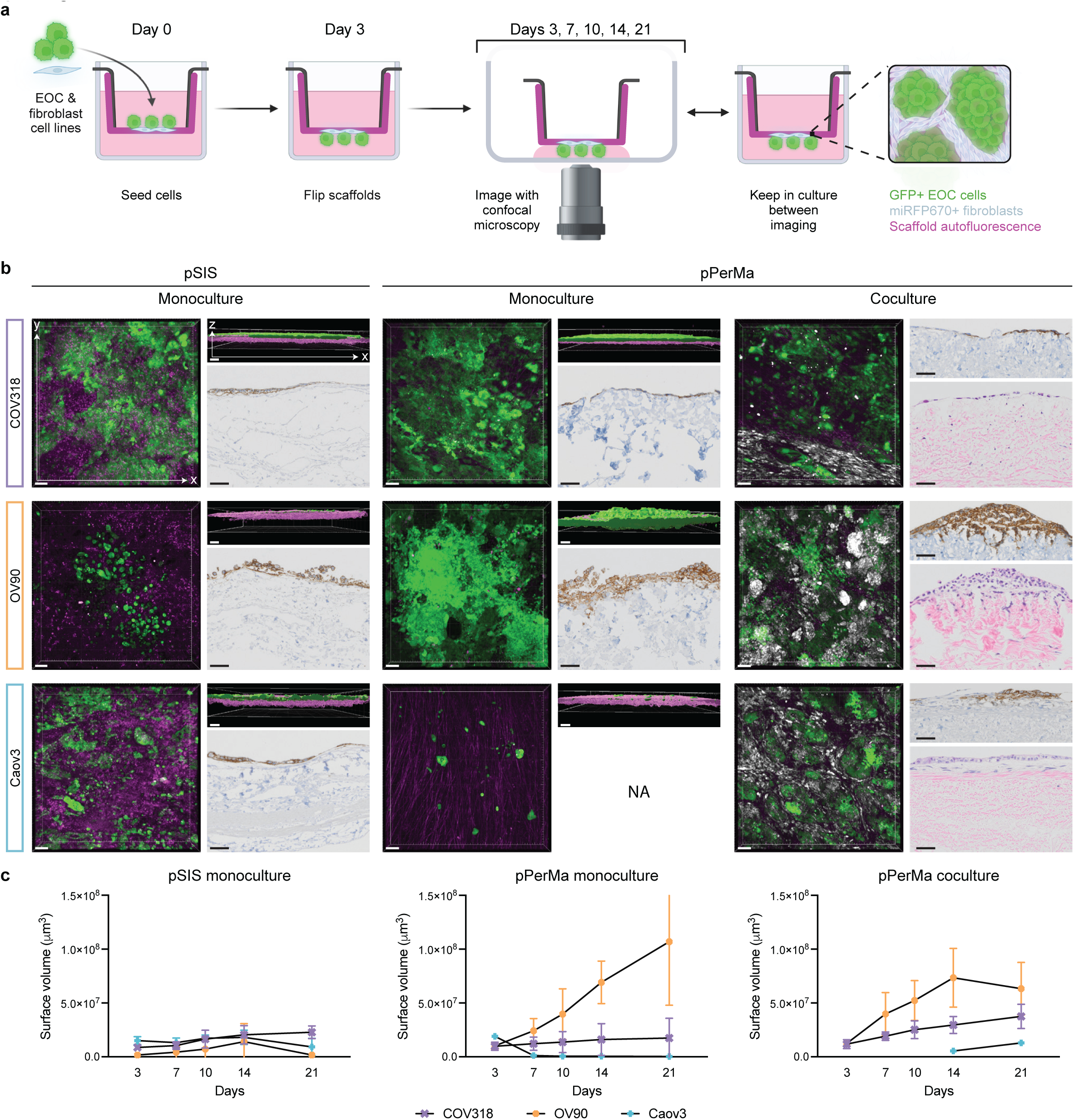
3D cell cultures are established with the porcine dECM scaffolds. **a** Schematic illustration of the setup for cell culture. Scaffolds were mounted in a Transwell®-like setup and cells were seeded in the inner chamber. After three days, the scaffolds were flipped, allowing the cells to come in direct contact with the imaging surface, enabling confocal imaging. Scaffolds were kept in this reversed position in the culture plate between imaging. **b** Confocal images and representative images of histological staining of cell cultures on pSIS and pPerMa. Each row represents a cell line (COV318, OV90, and Caov3). COV318 and OV90 cells were seeded in a number of 25,000 cells/scaffold, while Caov3 cells were seeded at 400,000 cells/scaffold for monoculture and 200,000 cells/scaffold for coculture. For cocultures, BJ fibroblasts were seeded in a number of 12,500 cells/scaffold (COV318), 6,250 cells/scaffold (OV90), and 25,000 cells/scaffold (Caov3). Confocal images are 3D renderings of z-stacks, visualized in the x/y plane (left, quadratic images) and the x/z plane (right, rectangular images). Magenta represents tissue autofluorescence (ex = 561 nm, em = 575-625 nm), green represents GFP+ EOC cell lines (ex = 488 nm, em = 500-550 nm), and white represents miRFP670+ BJ fibroblasts (ex = 637 nm, em = 663-738 nm). Scale bars = 100 µm. Histological stainings were performed on a cross-section of the scaffolds, corresponding to the x/z plane in the confocal images. The monocultures are stained with EpCAM. Cocultures are stained with EpCAM (top) and hematoxylin & eosin (H&E; bottom). Scale bars = 50 µm. NA = not applicable, too few cells to stain. The experiment was performed two times with three replicates for each group. **c** Graphs showing the cell growth of COV318, OV90, and Caov3 cells on pSIS (monoculture) and pPerMa (monoculture and coculture with fibroblasts). The cell volume was estimated using the surfaces function in the IMARIS image analysis software. Data represent mean ± SD.

Both pSIS and pPerMa were cytocompatible and allowed cell growth. The three cell lines displayed distinct growth patterns. COV318 cells formed a monolayer on the surface of both pSIS and pPerMa, with similar growth kinetics (Fig. 6b, c, Supplementary Fig. 5a). There was no significant difference in cell volume between pSIS and pPerMa at any of the time points. OV90 cells formed superficial three-dimensional tumor-like structures, with an increased growth on pPerMa compared to pSIS (Fig. 6b, c, Supplementary Fig. 5b). The difference was statistically significant at all time points (p values 0.0001 day 3, 0.0032 day 7, 0.0101 day 10, 0.0004 day 14, and 0.0014 day 21 for unpaired, two-tailed t-test). Caov3 cells seeded in the same number as COV318 and OV90 did not grow, neither on the pSIS nor the pPerMa. When seeded at 400,000 cells/scaffold instead of 25,000, some cell growth was observed on pSIS but not on pPerMa (Fig. 6b, c, Supplementary Fig. 5c).

Several studies have found that cancer-associated fibroblasts (CAFs) promote peritoneal metastasis of EOC by contributing to adhesion and invasion (52–54). Therefore, we incorporated the human fibroblast cell line BJ in the 3D cell cultures on pPerMa. The BJ cell line was transduced with miRFP670, a far-red fluorophore, permitting simultaneous confocal imaging of GFP-positive tumor cells and fibroblasts. Interestingly, we found that Caov3 cells, which were unable to grow on pPerMa in monoculture, did grow in coculture with fibroblasts (Fig. 6b, c, Supplementary Fig. 6c). Moreover, the tumor cells formed clusters surrounded by fibroblasts, a pattern that is also seen *in vivo* (55). COV318 cells in coculture with fibroblasts grew as a monolayer separated from the fibroblasts, while OV90 cells grew interspersed with fibroblasts (Fig. 6b, Supplementary Fig. 6a, b).

### 3.7 The 3D cell cultures can be used for therapy sensitivity testing

To assess sensitivity to therapy in the 3D *in vitro* model, we developed a method to assess cell death using confocal microscopy. For the establishment of the technique, we treated the cells with carboplatin, a chemotherapeutical used in the treatment of EOC. To select carboplatin doses to be tested in the 3D system, we determined the EC50 value of carboplatin for the OV90 cell line in 2D culture by WST-1 assay (Supplementary Fig. 7a). The cells were treated for 72 hours with carboplatin at doses ranging from 1 µM to 3160 µM and an EC50 value of 45.60 µM for OV90 cells was determined. To validate the EC50 value, we performed a set of experiments to evaluate cell death by Annexin V/PI staining and flow cytometry in 2D cell culture (Supplementary Fig. 7b,c). The cells were treated for 72 hours with carboplatin doses ranging from 10 µM to 316 µM, including the EC50 dose of 45.6 µM. The cells displayed increasing cell death, defined as positive staining for Annexin V and/or PI (Q1-Q3) with increasing drug doses. The EC50 dose group had 54.5% cell death.

To assess cell death in 3D, we used confocal microscopy and stained for live, apoptotic, and necrotic markers (Supplementary Fig. 7d, e). OV90 cells cultured on pPerMa were treated with carboplatin at three different doses (10 µM, 50 µM, and 100 µM) for 72 hours. The scaffolds were stained with CytoCalcein Violet, labeling the cytoplasm of viable cells, Apopxin Green, a phosphatidylserine (PS) sensor labeling membranes of apoptotic cells, and 7-AAD, a DNA intercalator labeling cell nuclei in membrane-permeable necrotic cells. All dyes stained OV90 cells growing on pPerMa. The fluorescent signal could be quantified by the IMARIS image analysis software using the surfaces function for live cells and the spots function for apoptotic and necrotic cells (Supplementary Fig. 7e). However, the Apopxin Green dye also stained the scaffold itself and the staining of apoptotic cells was challenging to distinguish from background staining. For further therapy sensitivity testing, we chose to continue with only markers for live and necrotic cells.

To investigate the feasibility of our model system to assess sensitivity to novel cell-based therapies, we tested co-culture with CAR T cells. A CAR T cell product, directed towards tumor-associated glycoprotein 72 (TAG-72), was tested on TAG-72 positive OV90 cells. The CAR and Mock T cells were generated from PBMCs of three different healthy donors. The EOC cell lines were cultured on pPerMa for 8 days before treatment (Fig. 7a). Imaging was done on day seven to ensure growth on all scaffolds and assign them to treatment groups. On day eight, CellTrace Violet-stained T cells were seeded. After 24 hours, the scaffolds were flipped, stained with 7-AAD, and imaged with confocal microscopy. The volume of live tumor cells was estimated from the GFP+ signal using the surfaces function in the IMARIS image analysis software. The confocal images show the difference in live cells (green) and dead cells (magenta) in the treated and untreated groups (Fig. 7b). After 24 hours of treatment, there was a significant difference in live cell volume between OV90 cells treated with TAG-72 targeted CAR T cells (n = 9) compared to Mock T cells (n = 9, p = 0.0015) and untreated OV90 cells (n = 3, p = 0.0111; Fig. 7c). There was also a significant difference in live cell volume between OVCAR3 cells treated with MUC-16 targeted CAR T cells (n = 9) compared to Mock T cells (n = 9, p = 0.0149) and untreated (n = 3, p = 0.0054; Fig. 7d). In summary, our 3D *in vitro* model system on pPerMa can be used to assess sensitivity to traditional and novel therapies against EOC.

**Figure 7.**
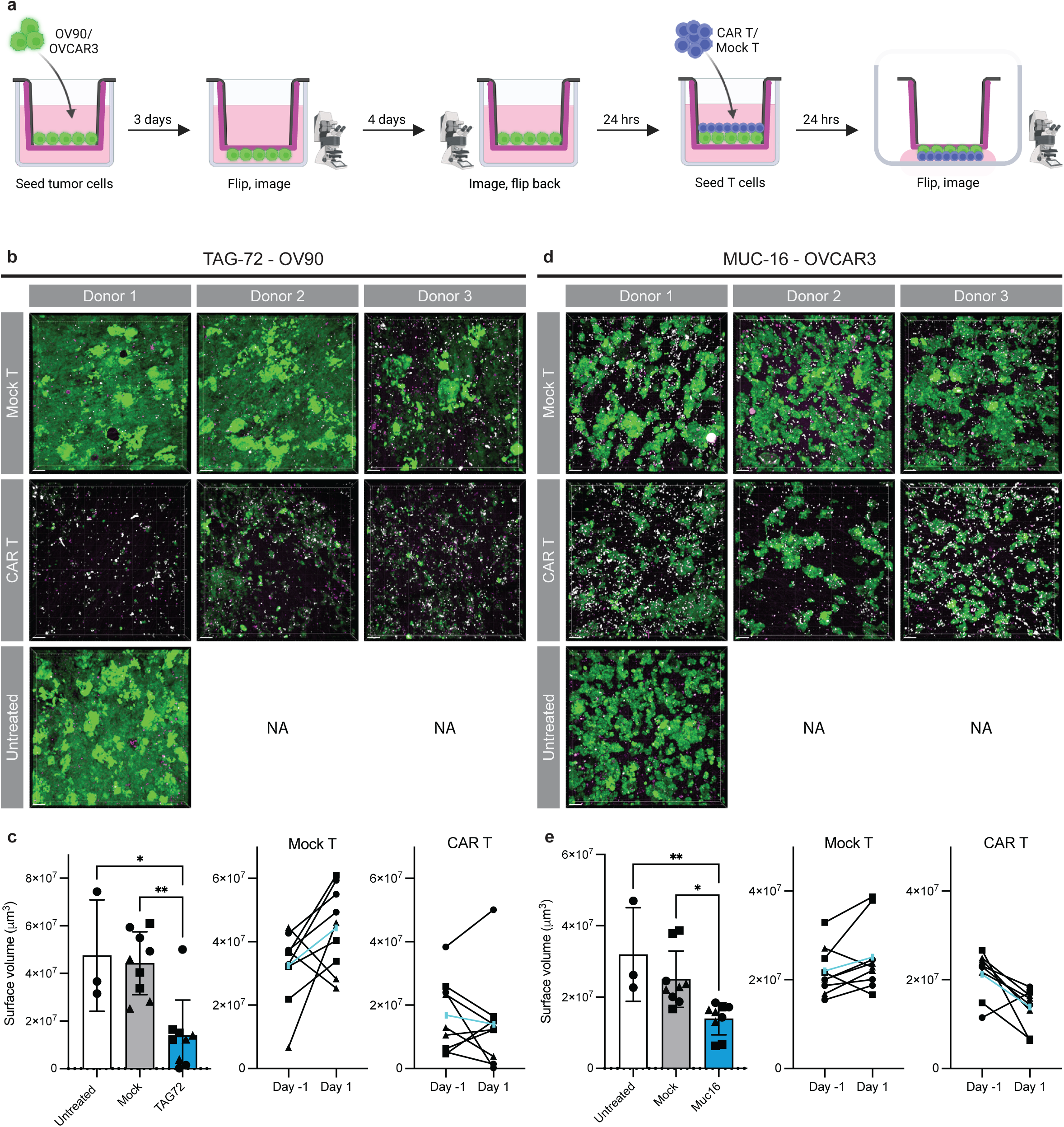
CAR T cell sensitivity is assessed in the 3D cell cultures using confocal microscopy. **a** Schematic illustration of the setup for treatment with CAR T or Mock T cells. The EOC cell lines OV90 and OVCAR3 were seeded on PerMa. OV90 cells were seeded at 25,000 cells/scaffold and OVCAR3 cells at 100,000 cells/scaffold. After three days, the scaffolds were flipped and imaged with confocal microscopy. After four more days, the scaffolds were imaged again and flipped back to the original position with the cells in the inner chamber. The next day, CAR T or Mock T cells were seeded on the scaffolds. After 24 hours, the scaffolds were flipped, stained with 7-AAD, and imaged with confocal microscopy. **b,d** Representative confocal images of OV90 cells treated with CAR T cells targeting TAG72 or Mock T cells (b), and OVCAR3 cells treated with CAR T cells targeting Muc16 or Mock T cells (d). CAR T and Mock T cells were derived from three different donors. The experiment was performed with three replicates in each group. Magenta represents 7-AAD (ex = 561 nm, em = 575-625 nm), green represents GFP+ EOC cell lines (ex = 488 nm, em = 500-550 nm), and white represents CellTraceTM Violet stained T cells (ex = 405 nm, em = 425-475 nm). The T cells that were adhered to the scaffold and/or tumor cells, did not detach when the scaffolds were flipped and can be seen in white on the confocal images. Scale bars = 100 µm. NA = not applicable. **c,e** Graphs showing the estimated cell volume of OV90 cells (c) and OVCAR3 cells (e) untreated and treated with Mock T cells or CAR T cells (left). Graphs showing the change in estimated cell volume from the day before treatment to 24 hours after treatment for Mock T cells (middle) and CAR T cells (right). The cell volume was estimated using the surfaces function in the IMARIS image analysis software. Data represent mean ± SD, * P < 0.05, ** P < 0.01. Different symbols in the graphs represent donor 1 (circle), donor 2 (square), and donor 3 (triangle).

## 4 Discussion

Peritoneal carcinomatosis is frequently associated with late-stage HGSOC and is a major therapeutic challenge. In this study, we established and characterized a 3D *in vitro* model of HGSOC peritoneal metastases utilizing a peritoneal dECM scaffold. Subsequently, we employed the model system to evaluate sensitivity to traditional chemotherapy and novel CAR T cell therapy.

Various advanced 3D *in vitro* models, ranging from patient-derived organoids (PDOs) to multicellular organotypics using artificial or tissue-derived hydrogels as scaffolds have been developed (27, 56, 57). These are limited in the recapitulation of the complex structure, biomolecular composition, and mechanical properties of the ovarian tumor ECM. Biological matrices made from the decellularization of tissues have proven to maintain the complexity of the native tissue ECM and have several advantages over hydrogels derived from one or a few ECM proteins (29). Herbert et al. utilized a porcine SIS (pSIS) scaffold to model peritoneal EOC metastases and demonstrated the utility of their model for studying tumor cell adhesion (31). However, differences in the ECM of the small intestine and the peritoneum may limit the SIS model’s representation of the peritoneum. To establish a model that more accurately mimics the ECM of the peritoneum, we developed a novel porcine peritoneal matrix (pPerMa). We seeded cells on both pSIS and pPerMa and noted significant differences in cellular proliferation rates between the two scaffolds (Fig. 6, Supplementary Figs. 5 and 6). Multiphoton microscopy and histological staining of major ECM components revealed only minor differences in collagen fiber arrangement, density and distribution of elastic fibers as well as mucins between pSIS and pPerMa (Fig. 3a-e). These variations are unlikely to fully account for the observed differences in cell growth on the two scaffolds. McKenzie et al. showed that higher stiffness in the environment leads to more focal adhesion formation, random migration, and migration directed by stiffness gradients (durotaxis) of tumor cells (58). This corroborates findings from studies reporting that tumor extracellular matrices often exhibit increased ECM rigidity (59, 60). Given the relatively minor variations in the rigidity of the pSIS, the pPerMa, and the hPerMa in our study, it is unlikely that the mechanical properties of the examined dECM scaffolds had a significant impact on the cultured cells. In contrast to common culture surfaces, such as polyethylene membranes that typically possess elastic moduli ranging from 2 GPa–2.7 GPa (61), the tested biological matrices closely mimic the soft structure of the native ECM. Consequently, they are a preferable option over artificial culture substrates.

Proteomic analysis showed that the pSIS and the pPerMa preserved the diverse composition of ECM proteins of the native tissues, including collagens, glycoproteins, and proteoglycans after decellularization (Fig. 5). The binding of ECM ligands to cell surface receptors, such as integrins, stimulates intracellular signaling pathways, facilitates cell adhesion, and regulates tumor cell migration (62, 63). Several of the glycoproteins and proteoglycans identified among the top 10 most abundant proteins in our samples, such as lumican (LUM), decorin (DCN), fibrinogen (FG), and biglycan (BGN), are dysregulated in cancer (63). We also identified important regulators of post-translational modification of ECM proteins, including transglutaminase (TGM) and cathepsin (CTS) (63) as abundant proteins, reflecting the diversity of ECM proteins in our samples. In contrast, many hydrogels consist of only one or a few different proteins. A recent study found that Matrigel®, commonly used to culture PDOs, is composed of more than 96% glycoproteins, 1% proteoglycans, and 0.4% collagen (64), which less faithfully replicates the characteristics of native tissue ECMs. Interestingly, we found differences in the matrisome profiles of the different dECM scaffolds (Fig. 5, Supplementary Fig. 4). First, the protein variety, or the total number of identified proteins, was comparable between the pPerMa and the hPerMa but was significantly higher in the pSIS (Supplementary Fig. 4). Second, the proportion of glycoproteins and proteoglycans was lowest in the pSIS but comparable between the pPerMa and PerMa from ovarian cancer patients (ocPerMa) (Fig. 5). These proteomic differences could influence the cell-matrix interactions in our 3D models. Given that both the pSIS and the pPerMa aim to mimic the ECM of native tissues, differences in their structure, composition, and mechanical properties underscore the importance of carefully selecting the appropriate tissue for model establishment.

We propose a porcine-derived PerMa as a suitable model due to its similarity to the hPerMa, its availability, and its reproducibility. Principal component analysis (PCA) confirmed high similarity between biological replicates in all groups, indicating little batch-to-batch variation, which is important to ensure experimental reproducibility. The comparison of the pPerMa and the hPerMa revealed similar organization and abundance of major ECM proteins (Fig. 3) and comparable compositions of core matrisome and matrisome-associated proteins (Fig. 5). We recognize the potential for proteomic disparities between the pPerMa and the hPerMa that our analysis did not reveal due to the challenges of directly comparing proteins from different species. As pointed out above, the rheological analysis uncovered differences in the viscoelastic properties of the pPerMa and the hPerMa. However, the differences are minor compared to other cell culture substrates. We confirmed the advantages of using porcine material, including the availability of tissue, the low batch-to-batch variation in the structure mechanical properties and biomolecular composition also reported in other publications using porcine material (34, 64, 65), in addition to pigs’ anatomic resemblance to humans (66).

Overall, the pPerMa can serve as a relevant biological matrix to study peritoneal metastases. Nevertheless, there are potential limitations of our model when employing animal tissues to model human disease. To better replicate human biology, the human PerMa established in this study could form the basis for the development of 3D models in future studies using primary patient cells on matched ocPerMa from the same patient.

We established 3D cell cultures by incorporating different cell lines in mono- and co-cultures. Cell lines offer advantages over primary cells as they are readily available and easy to use. We utilized cancer cell lines transduced to express GFP along with a fibroblast cell line transduced to express miRFP670. This enabled us to follow growth with confocal microscopy and study the interaction between the two cell types (Fig. 6, Supplementary Figs. 5 and 6). In our study, we observed that fibroblasts migrated deep into the submesothelial stroma while tumor cells remained in the more superficial layers, similar to what is seen in the native peritoneum and peritoneal metastases (8, 10) (Fig. 6). Further, in the coculture model of Caov3 cells and fibroblasts, the tumor cells formed clusters surrounded by fibroblasts in a pattern that is frequently seen *in vivo* (67) (Fig. 6). These findings may indicate that the PerMa plays a role in guiding cell migration and organization within tissues, which has been demonstrated for stem cells cultured on other dECM scaffolds (34). The pivotal ability to direct cell organization is crucial for integrating various cell types into a multicellular culture, thereby enhancing the representation of the TME. Incorporating additional cell types, such as endothelial, mesothelial, and immune cells is essential for improving the fidelity of our model. Malacrida et al. successfully replicated key elements of omental metastases in a coculture model consisting of mesothelial cells, fibroblasts, adipocytes, and HGSOC cells (68). Utilizing the PerMa as a scaffold could enable a similar reconstruction of peritoneal metastases.

Finally, we demonstrated the utility of our model for assessing sensitivity to traditional and novel cell-based therapies. Several drug screening platforms for HGSOC utilizing organoid technology have emerged (23, 25, 26), demonstrating potential in replicating patient responses to therapies and suitability for high-throughput drug screening. These systems commonly use quantitative assays that measure viability by detecting cellular metabolism via, for example, adenosine triphosphate (ATP). Our drug screening method differs technically from these platforms in several aspects, providing significant advantages. The custom-designed culture plate insert (referred to as a “crown”, Fig. 2) enabled us to set up the scaffolds in a Transwell®-like system with culture medium on both sides. The crown was adapted for confocal imaging. It can be downscaled to fit smaller culture plate wells, making it suitable for experiments with limited material availability. Our crown setup additionally allows for the simultaneous setup of migration and invasion assays, along with advanced imaging capabilities. With this configuration, we tracked the growth of the 3D cell cultures using confocal microscopy, both in standard cell culture conditions and during the therapy sensitivity assays and quantified live cells to assess the efficacy of chemotherapy and CAR T cell therapy (Figs. 6 and 7, Supplementary Figs. 5, 6 and 7). While frequently used bioluminescence assays are faster and easier to quantify, they do not provide the ability to examine cellular interaction, which is particularly advantageous for investigating novel cell-based immunotherapies. Our model is not a high-throughput screening platform but a highly valuable model to validate findings from other *in vitro* screening platforms to further explore how the interactions between cells and matrices could impact the therapeutic response of drugs and cell-based therapies.

In conclusion, we successfully established a 3D *in vitro* model of HGSOC peritoneal metastases using the pPerMa as a biological scaffold. The characterization demonstrated that the pPerMa closely recapitulates the ECM of the native peritoneum, making it a relevant platform for studying the unique TME of peritoneal metastases. Cell lines cultured on the PerMa show similar growth patterns to HGSOC *in vivo*. We demonstrated the utility of our model system for assessing sensitivity to traditional chemotherapy and novel cell-based immunotherapies. Overall, our model can serve as a valuable tool for the development of effective therapies for peritoneal carcinomatosis in HGSOC.

## Authors’ contributions

C.H.G., K.K., E.M., and L.B. conceived the study. C.H.G., K.K., and L.B. wrote the manuscript. C.H.G., A.L., and G.R.G.P. developed the decellularization protocol for peritoneum. C.H.G., G.N.S., A.L., and G.R.G.P. sampled and decellularized porcine small intestines and peritoneum. C.H.G., R.E., H.D., and D.E.C. sampled and decellularized human peritoneum. C.H.G., S.K., A.L., G.R.G.P., E.R. designed the 3D-printed cell culture plate inserts. B.D. performed histological staining. C.H.G. did multiphoton imaging. C.H.G., O.G., and K.L. performed multiphoton image analysis. C.B. did rheological assays and data analysis. C.H.G., M.S., and T.A.N. performed the protein extraction, mass spectrometry, and proteomics data analysis. C.H.G., A.L., G.R.G.P., E.R., and S.K. established protocols for 3D SIS and PerMa cell cultures and confocal imaging. C.H.G. G.N.S., and C.L. designed and performed the chemotherapy sensitivity assays. C.H.G., G.N.S., K.K., C.F., S.W., and P.G. designed and performed the CAR T cytotoxicity assays. K.K., E.M., and L.B. supervised the project. All authors have read and approved the final version of the manuscript.

## Acknowledgements

The authors gratefully acknowledge the patients who contributed to this study. Mass spectrometry-based proteomic analyses were performed by the Proteomics Core Facility, Department of Immunology, University of Oslo/Oslo University Hospital, which is supported by the Core Facilities program of the South-Eastern Norway Regional Health Authority. This core facility is also a member of the National Network of Advanced Proteomics Infrastructure (NAPI), which is funded by the Research Council of Norway INFRASTRUKTUR-program (project number: 295910). The authors acknowledge Viljar Helgestad Gjerde for assistance with the design of the 3D-printed staining ring, Marc Vaudel for guidance in proteomic analysis, Arild Holth for his assistance with the histopathological analysis, and Erdem Bozluolcay for assistance with the ECM structure analysis using TWOMBLI.

## SUPPLEMENTARY MATERIAL

**Supplementary Table 1.**

| **Antibody** | **Manufacturer** | **Cat. #** | **Clone** | **Host** | **Dilution** | **Antigen**<br>**retrieval** | **Positive**<br>**control** |
| --- | --- | --- | --- | --- | --- | --- | --- |
| Collagen I | Proteintech (Nordic Biosite) | 67288-1-Ig-20 | 1E9A7 | Mouse | 1:3000 | HpH (pH 9) | Normal human colon |
| Collagen IV | Invitrogen (Thermo Fisher) | PA1-28534 | Polyclonal | Rabbit | 1:400 | LpH (pH 6) | Normal human colon |
| EpCAM | Abcam | Ab20160 | AUA1 | Mouse | 1:200 | LpH (pH 6) | Human breast carcinoma |
| Fibronectin | NeoBiotechnologies | 2335-MSM2-P0 | TV-1 | Mouse | 1:100 | HpH (pH 9) | Human kidney |
| Laminin | Sigma Bio Sciences | L-8271 | LAM-89 | Mouse | 1:800 | LpH (pH 6) | Human kidney |

**Supplementary figure 1.**
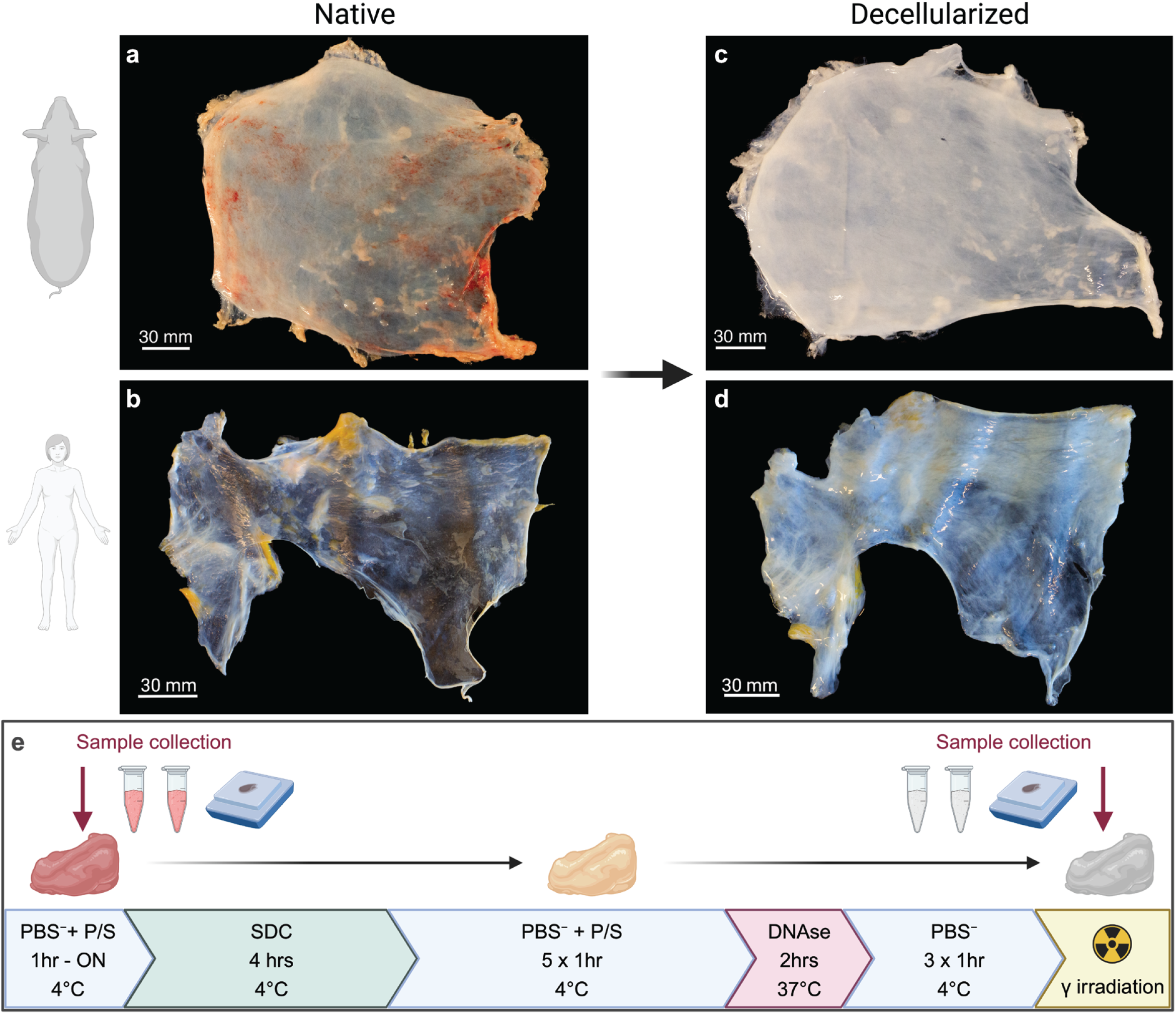
Photographs showing the native porcine (**a**) and human (**b**) peritoneum and the decellularized porcine (**c**) and human (**d**) PerMa. Scale bars 30 mm. **e** Schematic illustration of the protocol for decellularization of human and porcine peritoneum. Before decellularization, specimens were collected for formalin fixation and paraffin embedding (FFPE), and storage at −80 °C. Samples were washed in PBS with penicillin/streptomycin (P/S) for a minimum of 1 hour, maximum overnight (ON). Samples were incubated in sodium deoxycholate (SDC) for 4 hours at 4°C, followed by washing in PBS with P/S for a minimum of 5 x 1 hour at 4°C. Then, samples were incubated in DNAse solution for 2 hours at 37°C, followed by 3 x 1 hour washing in PBS. After decellularization, specimens were collected for FFPE and storage at −80°C. The scaffolds were then irradiated with gamma irradiation for sterilization.

**Supplementary figure 2.**
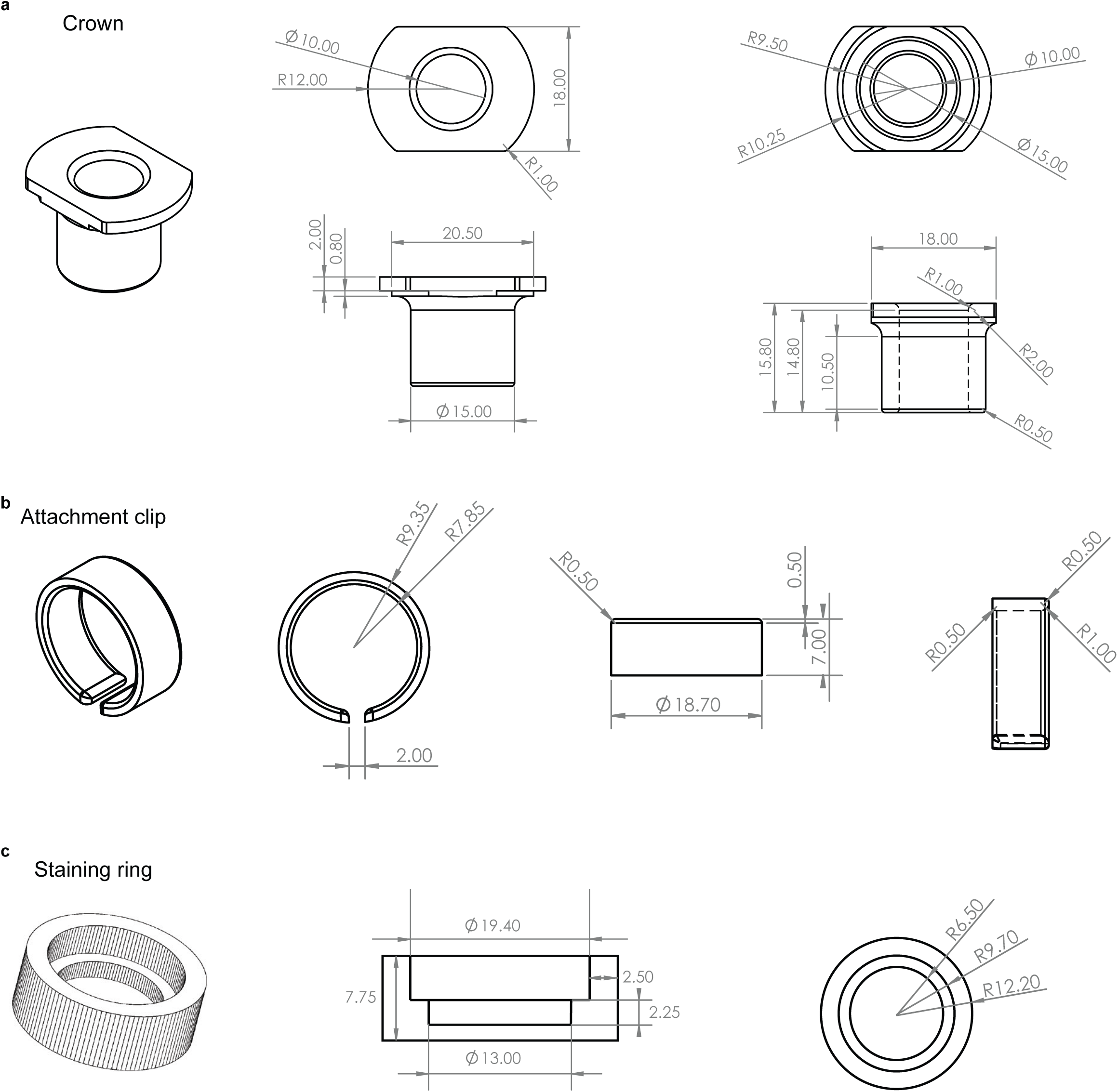
Sketches showing the dimensions of the cell culture crown (a), attachment clip (b), and staining ring (c).

**Supplementary figure 3.**
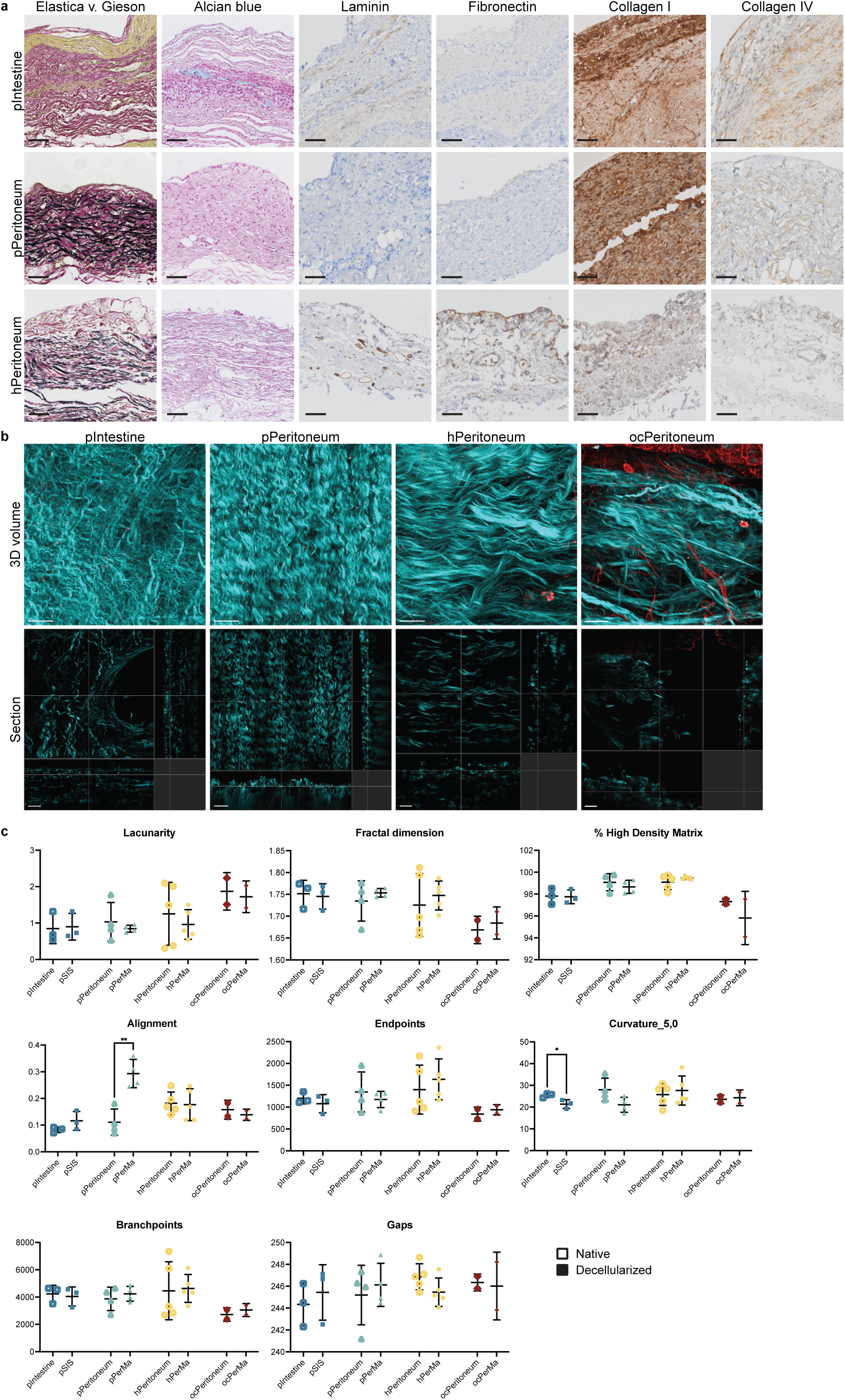
a. Representative images of Elastica v. Gieson and Alcian blue staining and immunohistochemical staining of laminin, fibronectin, collagen I, and collagen IV of pIntestine, pPeritoneum, and hPeritoneum. Scale bars 50 µm. **b** Representative second harmonic generation (SHG) microscopy images of pIntestine, pPeritoneum, hPeritoneum, and ocPeritoneum. 3D renderings (top) and single optical sections (bottom). Scale bars = 50 µm. **c** TWOMBLI analysis results. Data represent mean ± SD. *P < 0.05, **P < 0.01, unpaired two-tailed t-test performed for each pair of native/decellularized tissue. N = 3 for pIntestine/pSIS, n = 4 for pPeritoneum/pPerMa, n = 5 for hPeritoneum/hPerMa, n = 2 for ocPeritoneum/ocPerMa. Three representative images were captured and analyzed for each sample. Images were cropped to 250 µm (x) * 250 µm (y) * 10 µm (z) stacks. Four z stacks were generated per image area to include the top 40 µm from the surface.

**Supplementary figure 4.**
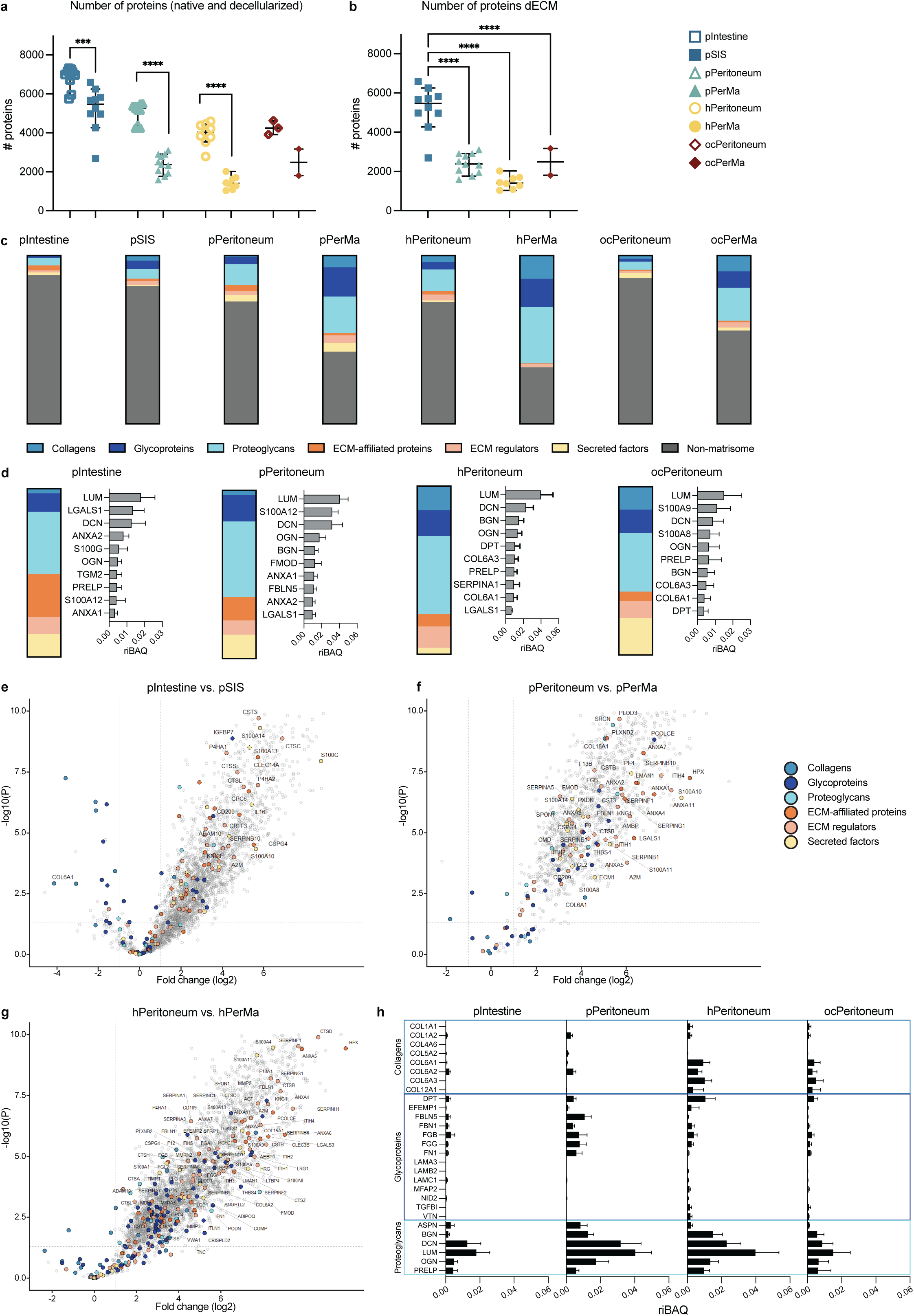
Proteomic analysis by liquid-chromatography mass spectrometry (LC-MS/MS). **a** Number of identified proteins in each group. Data represent mean ± SD. *** P < 0.001, **** P < 0.0001, unpaired two-tailed t-test performed for each pair of native/decellularized tissue. **b** Number of identified proteins in the dECM scaffolds. Data represent mean ± SD. **** P < 0.0001, one-way ANOVA with Tukey’s multiple comparison test. **c** The composition of matrisome and non-matrisome proteins in all groups. **d** The composition of total matrisome proteins and the ten most abundant matrisome proteins in pIntestine (n = 10), pPeritoneum (n = 11), hPeritoneum (n = 9), and ocPeritoneum (n = 3). The ratios of the top ten proteins are given as iBAQ intensity relative to the total iBAQ intensity (riBAQ) for each sample. Data represent mean ± SD. **e-g** Volcano plot of protein intensities in pIntestine (n = 10) vs. pSIS (n = 10; d), pPeritoneum (n = 11) vs. pPerMa (n = 11; e), and hPeritoneum (n = 9) vs. hPerMa (n = 8; f). Matrisome proteins with at least a 3-fold change (log2) are labeled. **h** Profile bar charts for pIntestine (n = 10), pPeritoneum (n = 11), hPeritoneum (n = 9), and ocPeritoneum (n = 3) of selected core matrisome proteins (n = 28). The proteins are sorted into collagens (top box), glycoproteins (middle box), and proteoglycans (bottom box). The ratios are given as relative iBAQ intensities. Data represent mean ± SD.

**Supplementary figure 5.**
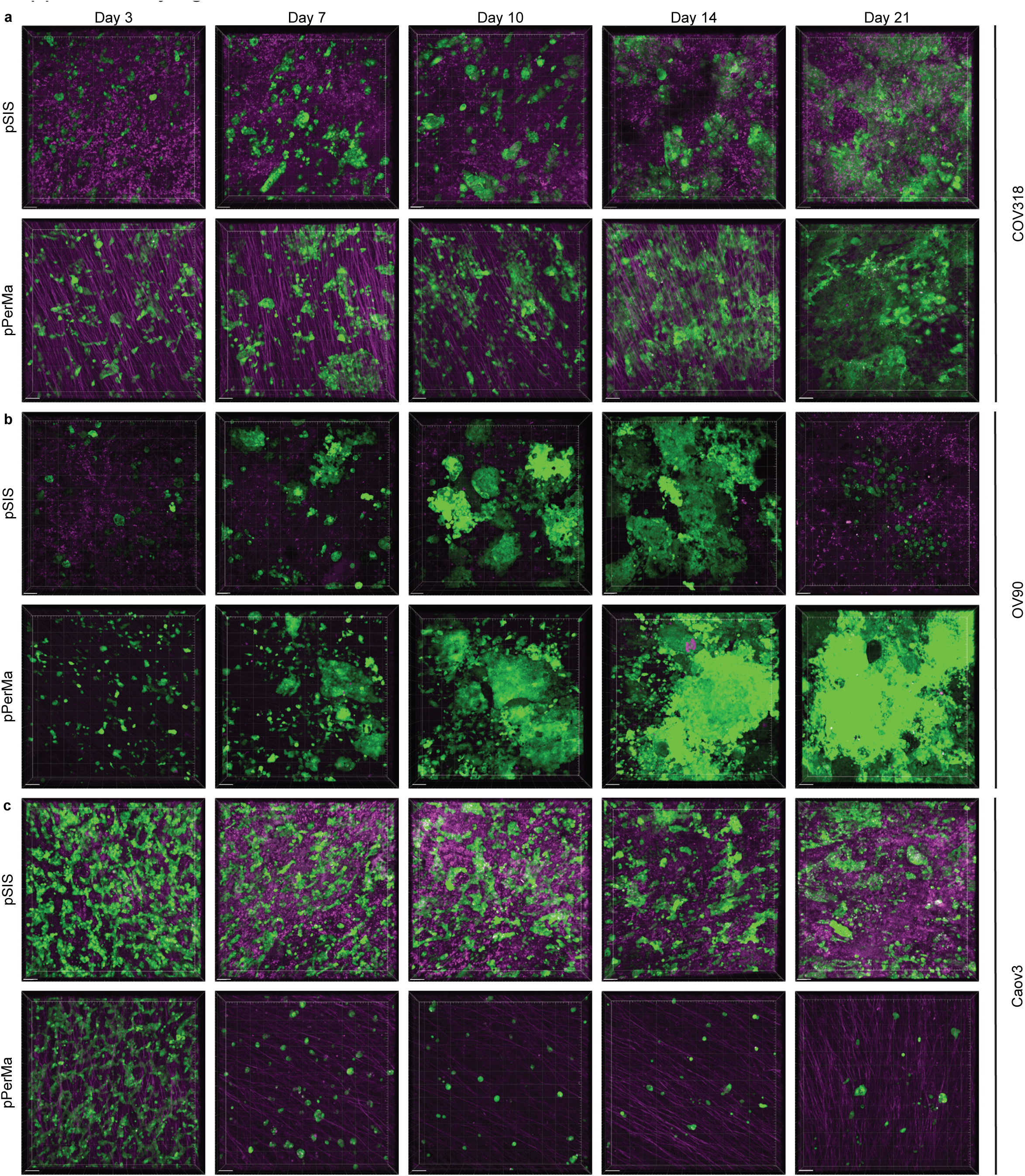
Representative confocal images of COV318 (**a**), OV90 (**b**), and Caov3 (**c**) on pSIS (top) and pPerMa (bottom) on days 3-21. COV318 and OV90 cells were seeded in a number of 25,000 cells/scaffold, while Caov3 cells were seeded at 400,000 cells/scaffold. The confocal images are 3D renderings of z-stacks, visualized in the x/y plane. Magenta represents tissue autofluorescence (ex = 561 nm, em = 575-625 nm) and green represents GFP+ EOC cell lines (ex = 488 nm, em = 500-550 nm). Scale bars = 100 µm.

**Supplementary figure 6.**
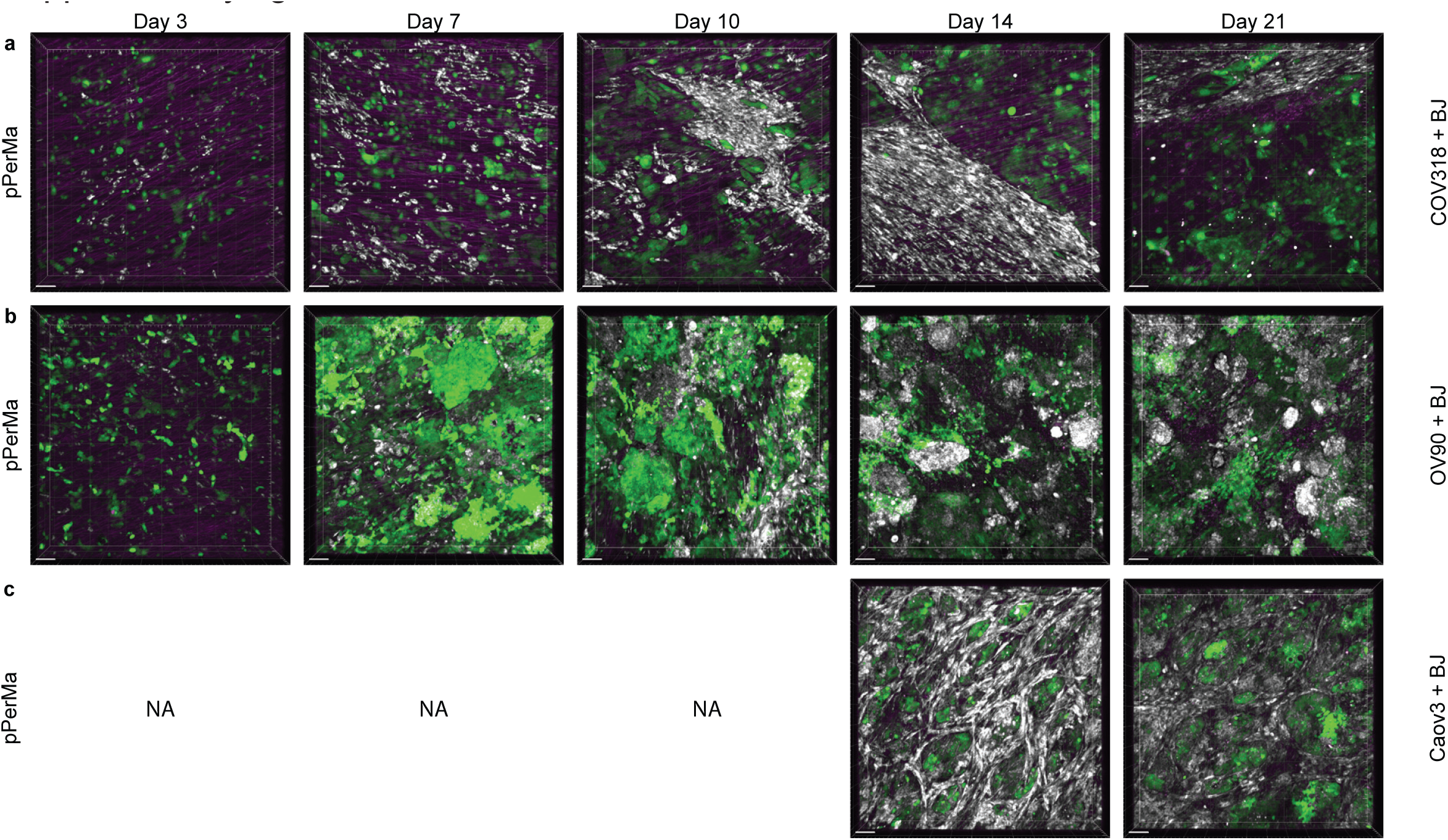
Representative confocal images of BJ fibroblasts in coculture with COV318 (**a**), OV90 (**b**), and Caov3 (**c**) on days 3-21. COV318 and OV90 cells were seeded in a number of 25,000 cells/scaffold and Caov3 cells at 200,000 cells/scaffold. For cocultures, BJ fibroblasts were seeded in a number of 12,500 cells/scaffold (COV318), 6,250 cells/scaffold (OV90), and 25,000 cells/scaffold (Caov3). The confocal images are 3D renderings of z-stacks, visualized in the x/y plane. Magenta represents tissue autofluorescence (ex = 561 nm, em = 575-625 nm), green represents GFP+ EOC cell lines (ex = 488 nm, em = 500-550 nm), and white represents miRFP670+ BJ fibroblasts (ex = 637 nm, em = 663-738 nm). Scale bars = 100 µm. NA = not applicable (only imaged on days 14 and 21).

**Supplementary figure 7.**
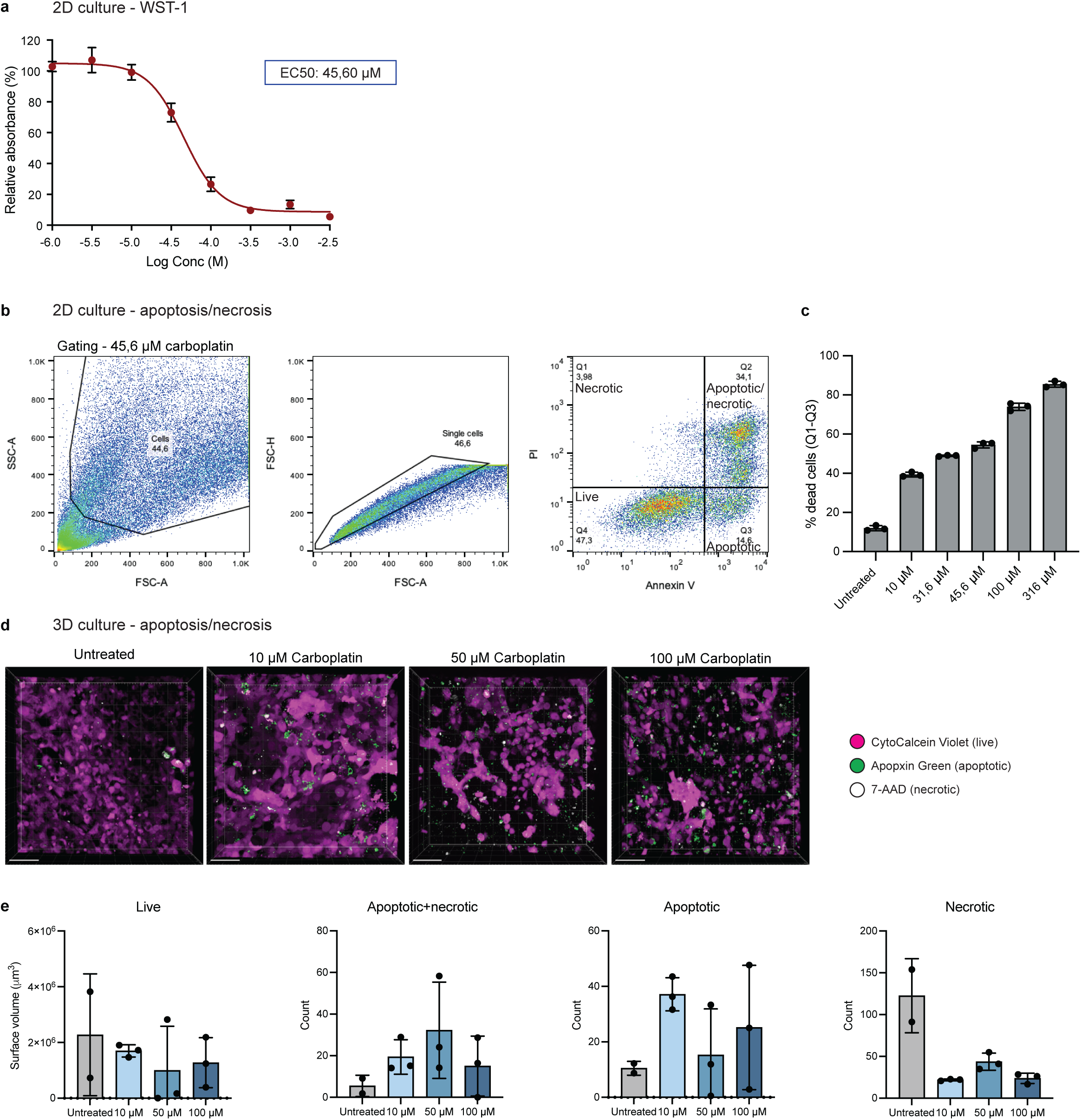
Carboplatin sensitivity for OV90 cells. **a** Dose-response curve for OV90 cells in 2D culture, treated with different doses of carboplatin. Y axis represents the absorbance of formazan in treated cells relative to control (%). X axis represents doses of carboplatin in moles/liter (M). The EC50 drug dose was determined to be 45.60 µM. **b,c** Response to carboplatin for OV90 cells in 2D culture, evaluated with Annexin V/PI staining and flow cytometry. **b** Example gating shown for one sample treated with 45.6 µM carboplatin. **c** Graph showing the percentage of dead cells for the different doses of carboplatin and untreated, calculated as the sum of Q1-Q3. Data represent mean ± SD. **d** Representative confocal images of OV90 cells cultured on pPerMa and treated with carboplatin at different doses (10 µM, 50 µM, and 100 µM). The confocal images are 3D renderings of z-stacks, visualized in the x/y plane. Magenta represents live cells stained with CytoCalcein Violet (ex = 405 nm, em = 425-475 nm), green represents apoptotic cells stained with Apopxin Green (ex = 488 nm, em = 500-550 nm), and white represents necrotic cells stained with 7-AAD (ex = 637 nm, em = 663-738 nm). **e** Graphs showing the surface volume (left graph) and cell count (three right graphs) of the markers for live, apoptotic, and necrotic cells for scaffolds treated with increasing doses of Carboplatin. Data represent mean ± SD.

